# ViDAS: Timely and Granular Estimation of Respiratory Virus Distributions in SARI Surveillance in Brazil

**DOI:** 10.64898/2026.11.25.26368960

**Authors:** João Flávio Andrade Silva, Guilherme P. Soares, Leonardo S. Bastos, Marcelo F. C. Gomes, Rafael Izbicki

**Affiliations:** Department of Statistics, Federal University of São Carlos, São Carlos, 13566-590, SP, Brazil; Institute of Mathematical Sciences and Computation, University of São Paulo, São Carlos, 13565-905, SP, Brazil; Fundação Oswaldo Cruz - Fiocruz, Presidência, Programa de Computação Científica - PROCC, Rio de Janeiro, 21040-900, RJ, Brazil; Instituto de Matemática Pura e Aplicada e Tecnologia - IMPA Tech, Rio de Janeiro, 20220-800, RJ, Brazil; Department of Statistical Science, University College London, London, WC1E 6BT, United Kingdom

**Keywords:** Severe Acute Respiratory Infections, Gradient Boosting Decision Trees, Viral proportion estimation, Nowcasting, Spatio-temporal epidemiology, Public Health Surveillance, Model interpretability

## Abstract

Effective monitoring of Severe Acute Respiratory Infections (SARI) is critical for public health but faces challenges such as reporting delays, incomplete information on negative test results, and data sparsity in specific regions and age groups. This study proposes a novel statistical framework to estimate the granular distribution of respiratory viruses among hospitalized SARI patients in Brazil. We employ Gradient Boosting Decision Trees (GBDT) to model viral proportions as conditional probabilities based on patient age, location, and time. A key methodological contribution is the development of SViDAS, a semi-parametric estimator that integrates viral probabilities over continuous lognormal age distributions, enabling estimation for arbitrary age intervals. Additionally, to address the “black box” limitations of machine learning, we introduce an interpretability strategy based on the estimator’s internal embedding space, which identifies relevant training instances to explain individual predictions. Using data from the Brazilian SIVEP-Gripe system (2022–2024), results demonstrate that our models reduce the Brier Score estimation error by approximately 40% for SARS-CoV-2 and 36% for RSV compared with the empirical aggregation benchmark. Furthermore, the reporting-delay analysis indicates that estimation error decreases sharply during the first few weeks and tends to stabilize after approximately 3 to 5 weeks for the main viruses, although the stabilization point varies by virus and geographical region. This approach provides a computationally efficient, high-resolution framework for timely surveillance of respiratory-virus distributions, including in municipalities with limited immediate data availability.

## 1. Introduction

Severe Acute Respiratory Infection (SARI) is a severe form of acute respiratory illness that can be caused by several infectious agents, including SARS-CoV-2, influenza viruses, and respiratory syncytial virus (RSV), and may progress to respiratory failure requiring hospitalization and intensive medical care [1].

In Brazil, universal surveillance of SARI was implemented in 2009 following the influenza A (H1N1) pandemic. Since 2019, SARI cases have been reported through the Influenza Epidemiological Surveillance Information System (SIVEP-Gripe), the official national system used to monitor the clinical and epidemiological profile of hospitalized cases and deaths, as well as circulating respiratory viruses [2, 3, 4]. Detailed epidemiological records are made publicly available and updated weekly by the Brazilian Ministry of Health [4, 3]. Although these data provide an important basis for respiratory-virus surveillance, obtaining timely estimates at fine spatial and demographic resolutions remains challenging. Viral circulation can vary substantially across locations, age groups, and epidemiological periods, while empirical estimates often require some degree of aggregation to ensure sufficient observations. Consequently, relevant local and demographic heterogeneity may be obscured when surveillance information is summarized over broader populations.

Our objective is to complement existing surveillance systems by providing timely estimates of the distribution of respiratory viruses among virus-positive SARI cases at a finer resolution, particularly across municipalities and patient ages. Rather than relying on predefined demographic and geo-graphical groups, the proposed framework models viral proportions as conditional probabilities based on location, population size, patient age, and epidemiological week. This formulation enables continuous monitoring of specific epidemiological contexts while leveraging information available across the surveillance dataset, supporting public health professionals in identifying changes in viral circulation that may inform prevention and control decisions.

To illustrate the level of granularity targeted by the proposed framework, Figure 1 presents estimated viral distributions across the 52 epidemiological weeks of 2024 for São Josédos Campos (state of São Paulo) and Uberlândia (state of Minas Gerais), comparing patients aged 1 and 65 years. The example illustrates substantial variation across both municipalities and age profiles that would be obscured under broader aggregation.

**Figure 1:**
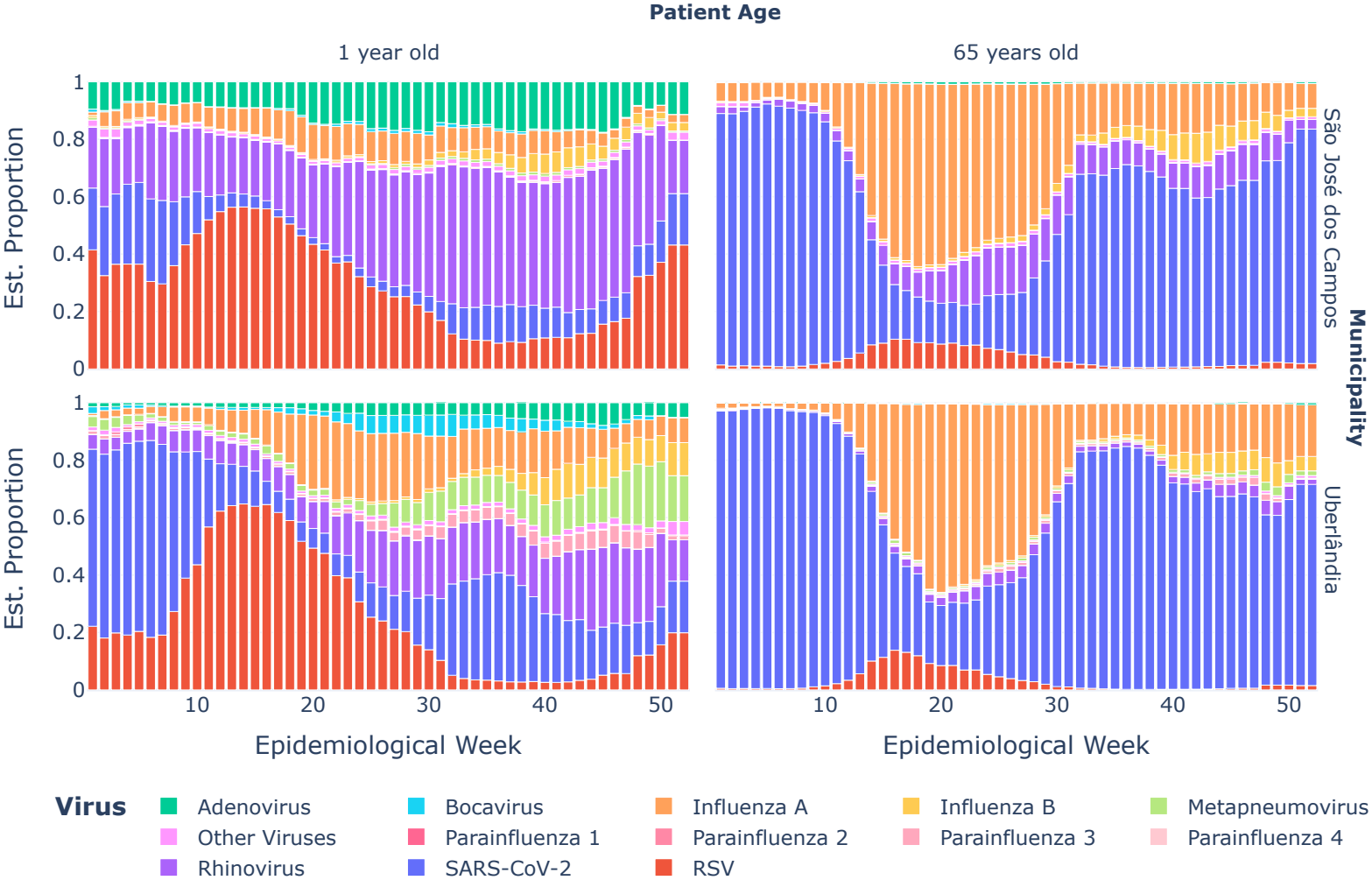
Estimated distribution of respiratory viruses over the 52 epidemiological weeks of 2024 as inferred by our method, stratified by municipality (São Josédos Campos and Uberlândia) and patient age (1 and 65 years). The panels reveal distinct, municipality-specific viral profiles across age groups: RSV and Rhinovirus predominate in infants (1 year), whereas SARS-CoV-2 and Influenza A are more prominent among the elderly (65 years).

The proposed methodology is implemented in a dedicated web application (https://joaoflavio.shinyapps.io/Virus_Monitor) for continuous visualization of the estimates [5]. We also provide the Python package PySRAG [6], which supports the processing and analysis of Brazilian SARI surveillance data and facilitates the reproducibility of the framework.

A preliminary version of this work was presented in [7]. The present study substantially extends it by introducing the SViDAS estimator for arbitrary age intervals, expanding the empirical evaluation to reporting-delay and data-scarcity scenarios, and providing an embedding-space approach for interpreting individual predictions.

### 1.1. Related work

#### Surveillance Systems

Current monitoring initiatives differ significantly in scope and methodology. The InfoGripe system [8, 9] employs Bayesian frameworks to estimate SARI incidence counts, correcting for reporting delays. However, its estimation of viral distribution—the proportion of cases attributed to specific pathogens—often relies on empirical aggregation over large geographical areas and extended time windows.

In contrast, the Instituto Todos pela Saúde (ITpS) [10] leverages data from a network of private laboratories to monitor pathogen circulation and compares these private positivity rates with SARI (SIVEP-Gripe) curves. However, two differences are particularly relevant for the specific goal of SARI surveillance. First, similar to the InfoGripe system, ITpS estimates are typically aggregated over large geographical areas (national, regional, or state levels), which obscures the heterogeneity of viral circulation at the municipality level. Second, the populations differ: SARI surveillance targets severe cases requiring hospitalization, whereas private laboratory data are based on individuals tested in partner laboratories, covering a broader clinical spectrum from mild to severe respiratory infections. Consequently, the viral distribution observed in private laboratory testing may not accurately mirror the etiological profile of severe infections, as pathogens associated with milder diseases may occur at different rates than among hospitalized SARI cases.

Our work bridges this gap by providing a granular, patient-level estimator that relies exclusively on SARI notification cases. Unlike empirical aggregation, which obscures local heterogeneity [11], our approach allows for specific viral attribution even in municipalities with sparse data.

#### Spatiotemporal Modeling

To estimate disease risk in areas with sparse data, the literature has traditionally leveraged spatial and spatiotemporal dependence via model-based geostatistics [12, 13]. Bayesian hierarchical models and Generalized Linear Mixed Models (GLMMs) are frequently employed to smooth incidence rates, explicitly incorporating spatial dependence through neighborhood adjacency matrices or distance-based covariance functions [14, 15].

However, these methods often require prior specification of spatial structures and, depending on the formulation, explicit interaction terms. While efficient approximations like INLA [16] exist, they can be sensitive to modeling assumptions [17] and computationally demanding for high-dimensional, real-time surveillance systems.

Consequently, recent literature has explored Machine Learning (ML) alternatives [18, 19]. Our work adopts this paradigm by utilizing Gradient Boosting Decision Trees (GBDT) [20], treating geolocation (latitude and lon-gitude) as continuous covariates. This allows the estimator to adaptively partition the spatial domain—effectively learning data-driven neighborhoods— and to capture complex, non-linear interactions between location, time, and patient age without requiring explicit interaction terms.

#### Complementing Nowcasting Efforts

Finally, this work complements the extensive *nowcasting* literature [21, 8, 22, 23] by shifting the target from incidence counts to viral proportions. This shift allows for the characterization of the etiological landscape even when total case counts are still being consolidated.

Beyond estimation, we address the *black box* criticism often leveled at ML models in public health [24]. We introduce a model-specific interpretability strategy based on the estimator’s internal embedding space, identifying influential training instances to explain individual predictions. Furthermore, we extend the standard point-estimation framework by proposing SViDAS, a semi-parametric estimator that integrates viral probabilities over continuous age distributions, enabling estimation for arbitrary age intervals (e.g., 0–2 years) relevant to age-specific surveillance.

## 2. Methodology

This work proposes a statistical framework designed to provide granular and timely estimates of respiratory viral distributions. Rather than relying on raw aggregates, we model viral proportions as conditional probabilities estimated through a machine learning approach. This section describes the main challenges associated with the surveillance data, dataset composition, and statistical procedures used to generate the proposed estimates.

### 2.1. Surveillance Challenges

To obtain timely and granular estimates of viral proportions, several challenges must be addressed:

1. **Information Delays:** Delays in data availability arise from the time required for information to pass through several stages, from an individual’s first symptoms perception to system reporting with virus identification. These stages include seeking medical care, undergoing testing, obtaining results, and entering the results into the surveillance system [8].
2. **Incomplete Data on Negative Tests:** The database records a virus only when the test result is positive, except for influenza. Consequently, the data do not distinguish individuals with negative results from those who were not tested for a given virus.
3. **Data Sparsity in Some Regions:** Information availability varies substantially across geographical areas and demographic groups. For example, in São José dos Campos, as shown in Figure 1, obtaining sufficient observations for 1-year-old patients is highly unlikely. Such sparsity may result from factors including lower population density, limited healthcare resources, or infrequent reporting within specific demographic groups.
4. **Computational Efficiency:** The models must be updated weekly and deployed in a web application with limited memory resources (e.g., 1 GB RAM), making computational efficiency an important methodological constraint.

To address incomplete information on negative tests (2), we estimate the proportion of each viral agent among SARI patients with at least one positive viral test. This estimand should therefore be interpreted as the viral composition among detected positive cases, rather than as the absolute incidence or positivity rate of each virus in the underlying SARI population.

This proportional formulation may also reduce sensitivity to reporting delays (1) when delays affect viral agents approximately uniformly, such that the relative composition of positive cases is not systematically distorted. In Section 3, we empirically assess this assumption by comparing estimates obtained under different reporting lags with the consolidated data.

Data sparsity (3) is commonly addressed by aggregating observations across regions or age groups, which inevitably reduces granularity. Instead, our methodology estimates the probability of observing a specific virus conditional on relevant covariates. A probabilistic multiclass classification algorithm leverages patterns across the full dataset to generate contextualized estimates for specific combinations of location, population size, patient age, and epidemiological week, including settings with sparse observations.

Finally, computational efficiency (4) is addressed through the choice and configuration of the underlying learning algorithm, enabling efficient model training and prediction under the operational constraints of the surveillance application. Detailed model configuration and training procedures are provided in Section S1 of the Supplementary Material.

### 2.2. Data

Since its implementation in Brazil in 2009, SARI universal surveillance has required the collection and reporting of all hospitalized cases and/or deaths due to respiratory viruses [2, 4]. The current Brazilian Ministry of Health’s definition characterizes SARI as an influenza-like illness (ILI) presenting with at least two of the following signs and symptoms: fever, chills, sore throat, headache, cough, runny nose, or olfactory or gustatory disturbances. In addition, the individual must exhibit one or more of the following: shortness of breath/respiratory discomfort, persistent chest pressure or pain, oxygen saturation below 95% in ambient air, or bluish coloration (cyanosis) of the lips or face [2].

Although the SARI criterion established by the Brazilian Ministry of Health is fundamental for national surveillance, its formulation was historically directed toward Influenza monitoring and was more recently adapted for the COVID-19 pandemic. The growing relevance of other respiratory pathogens, such as Respiratory Syncytial Virus (RSV), has led to discussions in global health organizations about broadening case definitions to improve their detection [25, 26].

Fever, a central component of the traditional criterion, is an inconsistent clinical sign in RSV infections, often being absent or blunted in vulnerable populations such as the elderly [27, 28], while also being a non-specific symptom with multiple causes in young children [29]. To enhance surveillance sensitivity for RSV and other respiratory viruses while maintaining comparability across seasonal and epidemiological periods, this study adopts a modified criterion that excludes the mandatory presence of fever. This approach aligns with the methodology employed by the InfoGripe surveillance system (Fiocruz), in accordance with updated Ministry of Health guidelines [30]. Accordingly, the datasets were filtered using the following logical rule: `(cough OR sore throat) AND (dyspnea OR oxygen saturation < 95% OR difficulty breathing) AND (hospitalized OR deceased)`. This strategy prioritizes respiratory and severity signs that are more consistent across different etiological agents.

Data were retrieved from the Brazilian Ministry of Health’s Open Data Portal [31]. To incorporate spatial and demographic dimensions, the records were enriched with geographical coordinates from the IBGE Selected Localities Register [32] and municipality population sizes based on the 2022 Demographic Census [33].

Formally, the dataset is denoted by 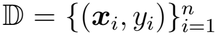, where ***x***_*i*_ contains information about the *i*-th individual, including the epidemiological week of symptom onset, patient age in years (computed from the reported age value and unit), geographical coordinates (latitude and longitude of the city, state, and region), and the population size of the notifying municipality. These covariates reflect relevant epidemiological dynamics: respiratory-virus prevalence varies substantially across age groups, while geographical and temporal information is particularly important in a country of continental dimensions such as Brazil, where viral circulation may differ markedly between regions at the same time. These choices are consistent with studies identifying age as a critical risk factor and highlighting regional disparities in SARI incidence [11, 34].

The response variable, *y*_*i*_, indicates the virus for which the patient’s test result was positive: SARS-CoV-2, Influenza A/B Virus, Respiratory Syncytial Virus, Human Parainfluenza Virus 1/2/3/4, Human Adenovirus, Human Metapneumovirus, Human Bocavirus, Human Rhinovirus, and/or Others (other viruses or unspecified). The dataset includes only patients who tested positive for at least one virus. In cases of viral co-detection, the patient record was expanded into multiple observations, one for each detected virus. A sample representation of the resulting dataset D is provided in Supplementary Table S1.

### 2.3. Statistical Framework

This subsection details the statistical methodology employed to estimate the conditional distribution of viral infections. We first define the multiclass classification framework used to estimate the class-conditional probability distribution over all viruses given a patient’s covariate profile. Subsequently, we extend this framework to estimate the marginal virus distribution over specific patient age intervals, employing a semiparametric integration strategy. Finally, we describe the embedding-based approach used to interpret individual estimator predictions by identifying influential training instances.

#### 2.3.1 Virus distribution estimator

Our goal is to estimate the proportion of patients who test positive for a specific virus among positive patients, conditioned on ***x***. We approach this task as a multiclass classification problem, where the variable *Y* can take values *y* representing different viruses being considered. The proportion is modeled by the conditional probability, denoted as ℙ_*Y*_ (*y*|***x***), which indicates the likelihood of a patient testing positive for a specific virus given the value of ***x***. To estimate this probability, we employ a probabilistic classifier, denoted as *ĝ*(*·*), obtained based on the data in D.

For the function *ĝ*(*·*), we chose the Gradient Boosting Machine (GBM) estimator based on decision trees, widely known as Gradient Boosting Decision Trees (GBDT; see Section S1 of the Supplementary Material for detailed model configuration and training strategy). In a multi-class problem like predicting viruses, the GBDT constructs a separate additive model for each possible virus class *k*. Each of these models produces a raw score (or logit), which represents the evidence for that specific class. The estimated score for a given virus *k*, which we denote as *ĥ*_*k*_ (***x***), is calculated as the sum of the predictions from all the trees built for that class:

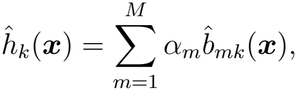

where *M* is the total number of trees in the ensemble for each class, 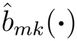 is the *m*-th decision tree assigned to the *k*-th virus class, and *α*_*m*_ is the learning rate, adjusting each tree’s contribution [35].

These raw scores, *ĥ*_*k*_ (***x***), are not probabilities; they are unbounded real numbers. To convert the vector of scores [*ĥ*_1_(***x***), *ĥ*_2_(***x***), …, *ĥ*_*K*_ (***x***)] into a valid probability distribution, we use the *softmax function*. The final estimated probability for a specific virus *k*, denoted *ĝ*_*k*_(***x***), is given by substituting the additive estimator directly into the softmax equation:

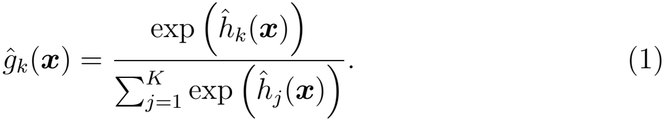

Here, the numerator is the exponentiated score for the virus class *k*, and the denominator is the sum of exponentiated scores across all possible virus classes *j* ∈ *{*1, …, *K}*. This transformation ensures that each probability *ĝ*_*k*_(***x***) is between 0 and 1 and that their sum across all classes equals 1, forming a proper probability distribution. The complete estimator, *ĝ*(***x***), is the function that outputs this full vector of probabilities. This estimator was trained using the multi-class logarithmic loss (multi-logloss), which is suitable for multiclass classification as it works in conjunction with the softmax function to ensure the predicted values from *ĝ*(***x***) approximate these valid class-conditional probabilities [36].

The choice of a tree-based estimator is due to its effective partitioning of the covariate space. It allows detailed analysis across geographical and temporal dimensions and interaction with other relevant variables, eliminating the need for predefined groups and offering a more flexible and precise data analysis approach. For a theoretical justification on the use of geographic coordinates over indicator variables for spatial partitioning in tree-based models, refer to Section S2 of the Supplementary Material.

This method was also chosen for its practical advantages: it allows for fast deployment due to low computational cost at prediction time and has low memory requirements for storing the estimator. Furthermore, GBDT performs implicit variable selection during the boosting process. The estimator’s structure also offers high interpretability; its tree-based nature supports global variable importance analysis, while its ensemble nature allows for the specific local prediction explanations (based on an internal embedding space) detailed in Section 2.3.3 and illustrated in Section 3.

#### 2.3.2. Estimating virus distribution in age intervals

The estimator *ĝ*(***x***) provides estimates of the virus distribution for a patient with SARI given a specific age, location, and epidemiological week of symptom onset. However, epidemiological surveillance often requires estimates for age groups rather than for a single patient age, such as infants under 1 year of age or adults between 60 and 70 years old. To obtain estimates for an age interval [*a, b*], we proceed in two steps: first, we estimate the conditional distribution of patient age given the remaining covariates; second, we integrate the pointwise virus estimator *ĝ*(***x***) over this estimated age distribution. In our implementation, patient age is modeled conditionally using a lognormal distribution whose parameters are estimated through GBDT models.

The resulting estimator is

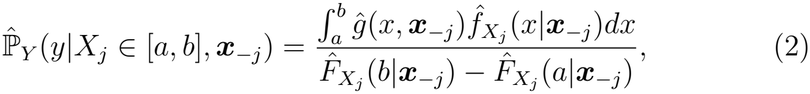

where 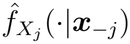 and 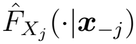 denote the estimated conditional density and cumulative distribution functions of patient age, respectively. This estimator represents the average predicted virus distribution over the age interval [*a, b*], weighted by the likelihood of observing each age within that interval conditional on the remaining covariates. In practice, the integral is evaluated numerically using a probability-weighted midpoint approximation. The complete derivation of the estimator, the GBDT-based estimation of the conditional age distribution, and the numerical integration procedure are provided in Section S4 of the Supplementary Material.

#### 2.3.3. Explaining Individual Predictions via Embedding Space Analysis

The definition of a neighborhood is an important component of local interpretation methods, including how it accounts for the geometry of the feature domain [37]. For gradient-boosted trees, the fitted ensemble itself can provide a representation for defining local groups. For example, LoBoost uses sequences of visited leaves to construct groups for local conformal calibration [38]. Here, we define similarity through an embedding constructed from the weighted outputs of the fitted trees. We then identify neighboring training observations to provide an example-based interpretation of individual predictions, including predictions for hypothetical cases in data-scarce contexts.

The similarity between any two observations is defined by their representation within the Gradient Boosting Decision Tree (1). For a given observation ***x***, its estimated embedding vector 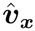 is constructed from the weighted outputs of each tree in the ensemble:

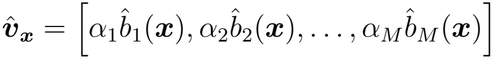

where 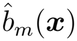 is the prediction from the *m*-th tree and *α*_*m*_ is its corresponding weight (learning rate). The similarity between a new case and instances in the training data is then quantified by calculating the Euclidean distance between their respective embedding vectors. This technique allows us to identify the training observations that are likely most influential in the estimator’s decision-making process for a specific prediction, thereby providing a clear, data-driven explanation for its output.

### 2.4. Summary of Proposed Estimators: The ViDAS Framework

Based on the statistical framework developed in the previous subsections, we formally define our proposed methodology, named **ViDAS (Viral Distribution estimation in Acute respiratory infection Surveillance)**. It is implemented through three distinct variants:

- **NViDAS (Nonparametric ViDAS):** As detailed in Section 2.3.1, this approach estimates the conditional probability of a patient testing positive for a specific virus, given the patient’s individual information.
- **ANViDAS (Aggregated Nonparametric ViDAS):** To obtain an estimate of the virus proportion for a specific municipality in each epidemiological week, the NViDAS predictive model is applied to each observed case in the database. Then, the average of the estimated probabilities is calculated for each week. This procedure results in a weekly estimate of the virus proportion that considers the empirical age distribution observed in the data for that week and municipality.
- **SViDAS (Semiparametric ViDAS):** As described in Section 2.3.2, this approach is used to estimate the virus proportion for a specific age range in a given location and epidemiological week. It employs the estimator combination methodology, which estimates the average virus probability for the age range of interest by weighting the predictions of the virus probability estimator by the estimated age distribution (log-normal) for the context. The result is a weekly estimate that reflects the average proportion for an age group, based on an estimated age distribution.

In summary, both ANViDAS and SViDAS are extensions of the base NViDAS model. While NViDAS operates at the individual level, estimating the probability for a patient with a specific profile (age, location, and week), ANViDAS and SViDAS aggregate these predictions to provide proportion estimates for populations defined by a municipality, an age range, and an epidemiological week.

## 3. Results

This section presents the results of the evaluation of the proposed methodology for monitoring viral infections in patients with Severe Acute Respiratory Infection in Brazil. Four main objectives are addressed:

1. To evaluate the performance of the methodology in estimating the distribution of respiratory viruses, comparing it with a standard approach and complete data (Consolidated Data).
2. To analyze the impact of reporting delays on the estimates.
3. To investigate the behavior of the methodology in data-scarcity scenarios.
4. To interpret the model, highlighting the importance of covariates and analyzing the influence of key predictor variables, particularly age and epidemiological week, on the predicted outcome.

The analyses were conducted using data released weekly between January 2022 and May 2025, referring to the years 2022, 2023, and 2024.

Based on the data released on May 5, 2025, SARS-CoV-2 was the pre-dominant virus during the 2022–2024 period, accounting for approximately 44.7% of the total positive cases. This was followed by Respiratory Syncytial Virus (RSV), Rhinovirus, and Influenza A, with 22.1%, 13.9%, and 8.7% of detections, respectively. Together, these four viral agents accounted for almost 90% of the occurrences in the analyzed period. The complete distribution across all viral categories is provided in Supplementary Table S2.

For the comparative analysis, we evaluate the proposed **ViDAS framework** (NViDAS, ANViDAS, and SViDAS, as defined in Section 2.4) against a standard model, referred to here as the **Naive Model**. This standard model is based on the empirical proportion of each virus by epidemiological week and State. Using only these variables, its main limitation is the inability to incorporate the granularity of age or municipality, producing a single estimate for each state per week.

The following subsections detail the analyses and discussions for each of the proposed objectives.

### 3.1. Performance Evaluation of the Proposed Methodology

This subsection evaluates the performance of the proposed models in estimating age-stratified viral proportions. Given this emphasis, the evaluation focuses on ANViDAS and SViDAS, which generate estimates for age intervals, a functionality not supported by NViDAS. Performance was assessed through two complementary approaches: a graphical case study and a systematic quantitative evaluation of estimation errors. In both analyses, the proposed models were compared with a Naive Model using the Consolidated Data, derived from the most complete database release available (May 5, 2025), as reference.

#### Graphical Analysis: Viral Dynamics and Model Performance

The graphical analysis focuses on the municipality of São Paulo during the first 26 epidemiological weeks of 2022. Estimates from ANViDAS, SViDAS, and the Naive Model were generated using data available on July 4, 2022 (epidemiological week 27), thereby simulating a nowcasting scenario. SARS-CoV-2 and RSV were selected due to their high incidence, and Figure 2 presents their estimated proportions for all ages and for two age groups with markedly different epidemiological profiles (*<*2 and 65+ years). Shaded areas represent the 95% confidence intervals for the Consolidated Data.

**Figure 2:**
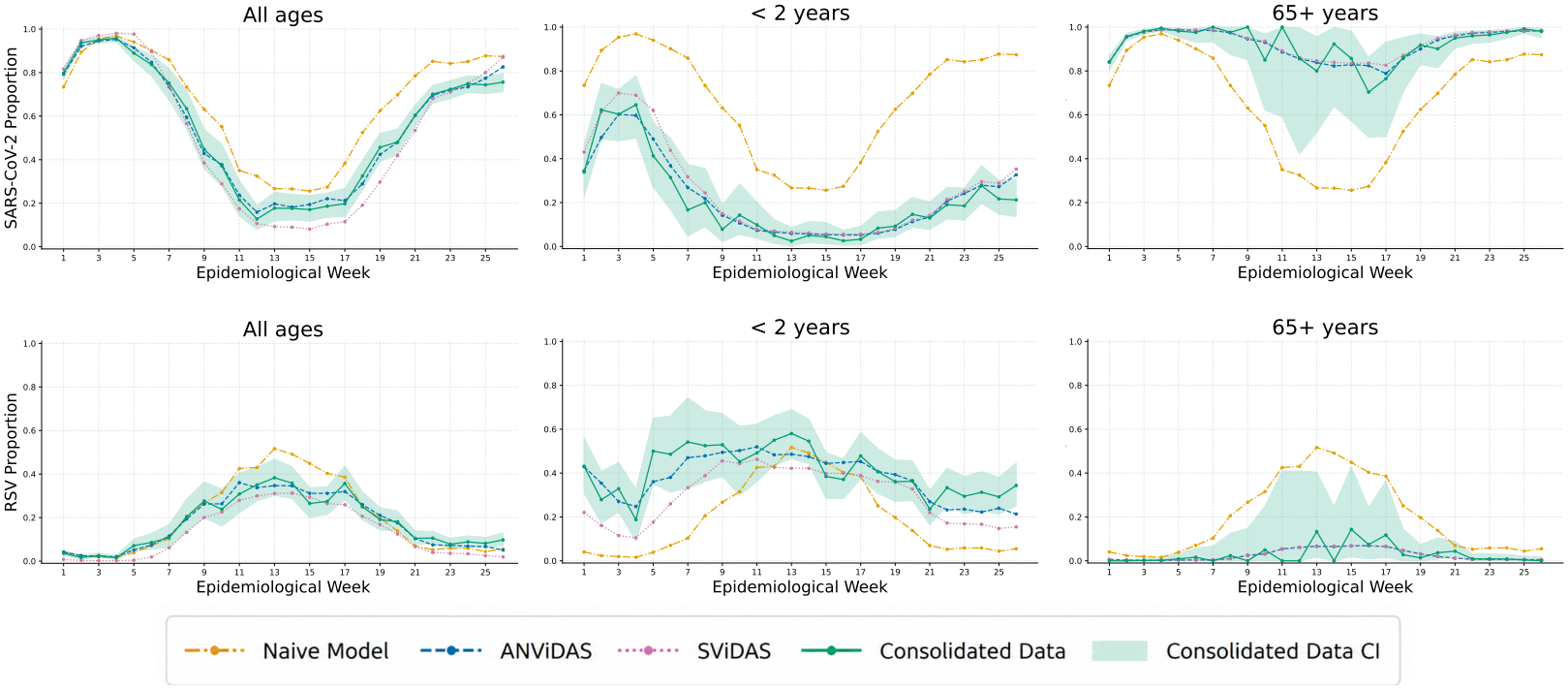
Estimated weekly proportions of SARS-CoV-2 and RSV among virus-positive SARI cases in São Paulo during the first 26 epidemiological weeks of 2022, shown for all ages, children under 2 years, and individuals aged 65 years or older. Estimates from **ANViDAS, SViDAS**, and the Naive Model are compared with the observed proportions from the Consolidated Data. Shaded areas represent the 95% confidence intervals for the Consolidated Data. The proposed models more closely follow the Consolidated Data trajectories across the selected age groups, whereas the Naive Model exhibits larger age-dependent deviations.

The curves reveal distinct temporal and demographic patterns for SARS-CoV-2 and RSV. Their proportions frequently exhibit opposite temporal trends, largely reflecting the proportional structure of the outcome and the predominance of these two viruses during the analyzed period, rather than direct viral competition.

ANViDAS and SViDAS consistently track the Consolidated Data more closely than the Naive Model. SViDAS, by modeling age continuously, produces smoother trajectories, particularly in strata with fewer observations, whereas ANViDAS more directly reflects weekly fluctuations in the empirical age distribution. The visual analysis also highlights systematic age-dependent errors in the Naive Model: it tends to overestimate RSV proportions in older individuals and underestimate them in young children, while the opposite pattern is observed for SARS-CoV-2. These results illustrate the importance of age stratification, with RSV predominating in the pediatric population and SARS-CoV-2 maintaining a higher proportion among older individuals during the analyzed period.

#### Quantitative Analysis: Validation of Estimation Errors

The quantitative evaluation complements the graphical analysis by simulating the operational use of the models. The models were retrained with each new data release between January 1, 2022, and May 5, 2025, and evaluated for epidemiological weeks within the 2022–2024 period. For each release, a new set of estimators was generated and used to predict the preceding 14-week window, focusing the evaluation on the period in which notification data are most incomplete and subject to updates. Performance was assessed using the Brier Score [39], which measures the mean squared difference between predicted probabilities and observed outcomes, with lower values indicating better probabilistic predictions. Estimates were compared against individual outcomes from the Consolidated Data.

The quantitative results, summarized in Table 1, corroborate the findings of the graphical analysis. ANViDAS and SViDAS consistently outperform the Naive Model across all 13 viruses, with substantial error reductions for the most incident agents. For SARS-CoV-2, for example, the average Brier Score is reduced by more than 40% compared with the Naive Model. ANVi-DAS and SViDAS show very similar performance, with ANViDAS having a marginal advantage in most cases, indicating that both approaches to incorporating age distribution—empirical and modeled, respectively—are effective. The lower standard deviations observed for the most prevalent viruses also indicate greater stability in predictive performance across evaluations.

**Table 1:** Mean Brier Score (and standard deviation) for each virus. The lowest error for each virus is highlighted in bold.

| **Virus** | **Naive Model** | **ANViDAS** | **SViDAS** |
|---|---|---|---|
|  | **Mean (SD)** | **Mean (SD)** | **Mean (SD)** |
| SARS-CoV-2 | 0.1703 (0.0840) | **0.0997** (0.0489) | 0.1011 (0.0499) |
| RSV | 0.1299 (0.0879) | **0.0825** (0.0691) | 0.0840 (0.0708) |
| Rhinovirus | 0.1252 (0.0837) | **0.1025** (0.0657) | 0.1032 (0.0661) |
| Influenza A | 0.1021 (0.0876) | **0.0811** (0.0608) | 0.0817 (0.0610) |
| Adenovirus | 0.0308 (0.0337) | **0.0287** (0.0314) | 0.0290 (0.0317) |
| Metapneumovirus | 0.0238 (0.0209) | **0.0226** (0.0200) | 0.0227 (0.0201) |
| Influenza B | 0.0211 (0.0357) | **0.0187** (0.0297) | 0.0189 (0.0302) |
| Other viruses | 0.0211 (0.0142) | **0.0200** (0.0136) | 0.0201 (0.0136) |
| Parainfluenza 3 | 0.0102 (0.0100) | **0.0096** (0.0097) | 0.0097 (0.0098) |
| Bocavirus | 0.0091 (0.0103) | **0.0085** (0.0099) | 0.0086 (0.0100) |
| Parainfluenza 1 | 0.0023 (0.0022) | **0.0021** (0.0021) | 0.0022 (0.0021) |
| Parainfluenza 4 | 0.0018 (0.0024) | **0.0016** (0.0023) | 0.0017 (0.0023) |
| Parainfluenza 2 | 0.0016 (0.0018) | **0.0015** (0.0016) | **0.0015** (0.0017) |

The age-stratified evaluation further shows that the advantages of the proposed models are particularly evident in age groups for which viral distributions differ substantially from the overall population pattern. This is especially apparent for RSV and SARS-CoV-2, for which the Naive Model exhibits systematic age-dependent errors that are substantially reduced by both ANViDAS and SViDAS. The proposed models also outperform the Naive Model across age groups for Rhinovirus and Influenza A, further supporting the importance of explicitly incorporating age into the estimation process.

Additional methodological details, complete age-stratified curves, detailed Brier Score results by age group, and a more comprehensive analysis and interpretation of the age-stratified performance are provided in Section S6 of the Supplementary Material.

### 3.2. Analysis of the Effect of Delays on Estimates

We evaluated the impact of reporting delays on the performance of the estimates generated by the proposed methodology. To this end, we conducted an experiment simulating the operational use of NViDAS over time, mimicking a nowcasting setting in which predictions are initially generated from partial information and subsequently updated as data are consolidated. The analysis was based on weekly data releases from January 2022 to May 2025, with a new NViDAS estimator trained for each release to reflect the information available at that specific point in time. Performance for each epidemiological week was evaluated as a function of the reporting lag, defined as the difference, in weeks, between the data release date and the epidemiological week for which the estimate was generated.

A consistent pattern was observed across Brazil and its five macro-regions: estimation error decreased sharply during the first few weeks and subsequently tended to stabilize. This rapid improvement may partly reflect the increasing volume of available data, but may also be influenced by the dynamics of laboratory confirmation, whereby cases with higher clinical probability may be confirmed earlier and additional results are progressively incorporated into the surveillance system. As the data are consolidated, the estimates become increasingly precise.

For the main viruses, the estimation error tends to stabilize after a lag of approximately 3 to 5 weeks, although the stabilization point varies by virus and geographical region. From approximately the third week onward, the error is already close to the lowest levels observed after further data consolidation in most cases. These findings support the applicability of NViDAS for timely epidemiological surveillance. Detailed results on the delay-analysis design, stabilization criterion, error trajectories, and regional patterns are provided in Section S6 of the Supplementary Material.

### 3.3 Performance in Data-Scarcity Scenarios

This subsection evaluates the performance of the proposed methodology in a data-scarcity scenario, a common challenge in epidemiological surveillance. To simulate this condition, we used the weekly datasets released between January 2022 and May 2025, covering the 2022–2024 period. For each data release, the models were trained on a small random subsample comprising 10% of the data and evaluated on the remaining 90%. The performance of NViDAS was compared with that of the Naive Model, using the Brier Score to assess the quality of the probabilistic estimates.

The results are summarized in Figure 3. NViDAS consistently outperforms the Naive Model, particularly for the most incident viruses. The advantage is most pronounced for SARS-CoV-2 and RSV, for which NViDAS achieves lower Brier Scores and lower variability than the Naive Model. Conversely, for viruses with very low incidence, such as the Parainfluenza group (PARA1–4) and Bocavirus (BOCA), the difference between the two models is less pronounced, with both exhibiting very low error scores.

**Figure 3:**
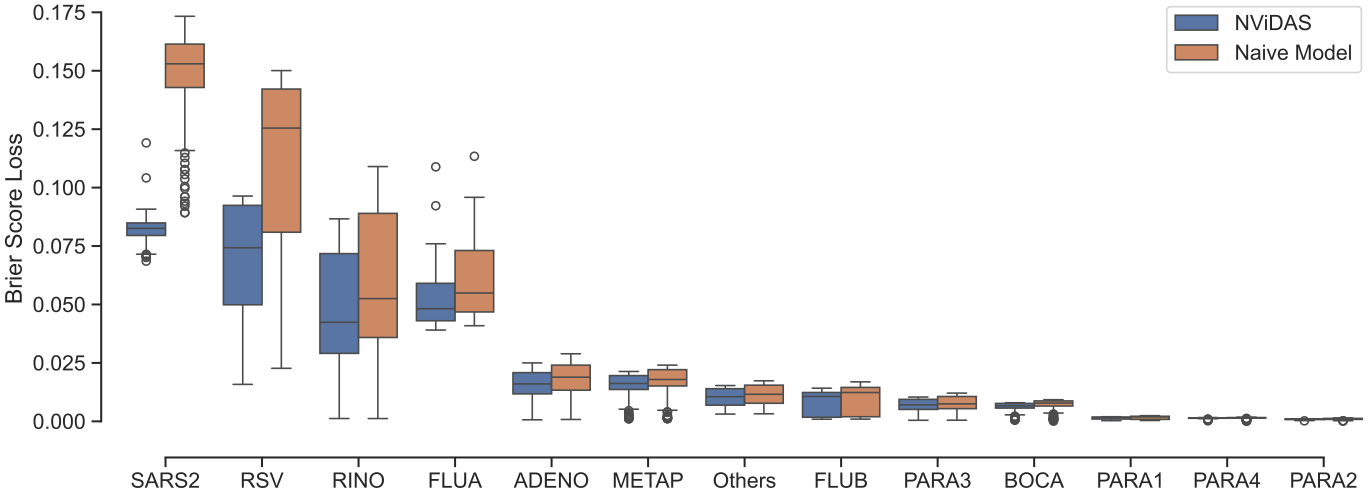
Performance comparison between **NViDAS** and the Naive Model in a data-scarcity scenario. The models were trained on 10% of the available data and evaluated on the remaining 90%. The boxplots show the distribution of Brier Scores for each virus, with lower scores indicating better performance.

Overall, this analysis supports the robustness of the proposed methodology under data-scarcity conditions. By leveraging relationships among covariates, NViDAS maintains superior predictive performance relative to the Naive Model even when trained on a limited amount of data, supporting its applicability to epidemiological surveillance in settings with sparse observations.

### 3.4. Interpretation and Explanation of the NViDAS Model

The global feature-importance analysis revealed a clear hierarchy among the predictor variables. Patient age emerged as the most influential predictor, followed by the epidemiological week of symptom onset. Geographical variables also exhibited a tiered importance according to their level of granularity, with city-level coordinates being more influential than state- and region-level coordinates. This pattern suggests that, in addition to demographic and temporal information, NViDAS captures relevant localized variation in viral transmission dynamics.

The analysis of the effects of age and epidemiological week further revealed distinct epidemiological patterns across the main respiratory viruses. For SARS-CoV-2, the model learned a strong positive relationship between age and predicted probability, with lower probabilities among infants and progressively higher probabilities among older individuals. In contrast, RSV exhibited a strong negative relationship with age, with the highest predicted probabilities among neonates and infants and a sharp decline during the first years of life. Rhinovirus showed a more complex, non-linear age pattern, with predicted probabilities peaking during childhood and gradually declining through adulthood. For Influenza A, epidemiological week had a particularly strong influence, characterized by pronounced seasonal peaks across age groups. Overall, these results indicate that the model learned not only the relevance of age and epidemiological week as individual predictors, but also virus-specific age and temporal patterns that reflect distinct epidemiological signatures.

We also investigated the interpretability of individual NViDAS predictions in settings where direct observations were unavailable. For this purpose, we applied the local interpretation method specifically proposed in this study and introduced in Section 2.3.3, which analyzes individual predictions through neighboring observations in the model’s embedding space. This analysis showed that, in data-scarcity settings, NViDAS bases its predictions on training profiles that are similar to the target observation in demographic, temporal, and geographical characteristics. The resulting explanations indicate that the model can draw on different sets of relevant observations depending on the patient’s age and epidemiological context, thereby supporting nuanced and age-appropriate predictions even when no direct observations are available for a given municipality.

Detailed results of the feature-importance analysis, the Partial Dependence Plot (PDP) and Individual Conditional Expectation (ICE) analyses, and the two case studies used to illustrate the proposed local interpretation method are provided in the Section S6 of the Supplementary Material.

## 4. Discussion

### Interpretation of Main Findings

This study presented a computational framework for estimating the granular distribution of respiratory viruses among SARI patients in Brazil. Our results indicate that modeling viral proportions as conditional probabilities, using GBDT, consistently outperforms the empirical aggregation benchmark. Specifically, the proposed models (ANViDAS and SViDAS) achieved a reduction in estimation error (Brier Score) of approximately 40% for SARS-CoV-2 and 36% for RSV compared with the benchmark.

These findings directly address the research objective of efficiently monitoring viral distributions across municipalities and age groups. Patient age emerged as the most influential predictor of viral distribution, followed by the epidemiological week. The model also captured distinct non-linear age-related patterns that are consistent with established epidemiological differences, including higher RSV proportions among infants and higher SARS-CoV-2 proportions among older SARI patients, with a sharp decline in predicted RSV probability during early childhood and a progressive increase in predicted SARS-CoV-2 probability with age.

### Comparison with Related Work and State-of-the-Art

Our approach represents a methodological shift from aggregate-level surveillance to patient-level estimation. Unlike empirical aggregation over broader geographical and demographic groups, our method preserves local heterogeneity. By treating geolocation as continuous covariates (latitude and lon-gitude), the model effectively learns data-driven neighborhoods without requiring pre-specified adjacency structures.

The ability of SViDAS to integrate viral probabilities over continuous lognormal age distributions constitutes a semi-parametric extension of the framework. This allows estimation in arbitrary age intervals (e.g., neonates vs. toddlers), offering a granularity that fixed-bin aggregation cannot match. Separately, to address the “black box” limitations often leveled at machine learning models, we introduced an interpretability strategy based on the estimator’s internal embedding space. By identifying training instances associated with a prediction, this approach provides data-driven explanations for the model’s outputs, including in data-scarce contexts.

### Methodological Considerations and Robustness

The choice of a GBDT architecture was justified by its ability to capture complex interactions between space, time, and demographics without the need for extensive feature engineering or predefined grouping. The decision to model the proportion of positive cases rather than total incidence was a strategic response to the absence of negative test records for non-influenza viruses in the SIVEP-Gripe database. Regarding reporting delays, this proportional formulation is expected to be less sensitive than absolute case counts when delays affect viral agents approximately uniformly, so that the relative composition of positive cases is not systematically distorted. The empirical delay analysis supports the practical applicability of this formulation: estimation error decreased sharply during the first few weeks and tended to stabilize after approximately 3 to 5 weeks for the main viruses, although the stabilization point varied by virus and geographical region.

The robustness of this approach is further supported by the data-scarcity analysis. Even when trained on only 10% of the available data, NViDAS maintained superior predictive performance relative to the Naive Model. In addition, the case study of Araporã showed that, even in the absence of local observations, the model could leverage the embedding space to identify similar training instances—geographically, temporally, and demographically related—to support predictions for the target profile. Together, these results support the applicability of the framework for timely surveillance in settings where data are still being consolidated or are locally sparse.

### Age-Density Specification in SViDAS

The SViDAS estimator uses a lognormal working model for the conditional distribution of patient age, chosen to enable computationally efficient estimation of viral proportions over arbitrary age intervals. In the empirical evaluation, SViDAS achieved performance very close to that of ANViDAS, which aggregates predictions from the same virus-probability model using the empirical age distribution. This close agreement suggests that, for the settings and age intervals evaluated in this study, the lognormal specification provides an adequate practical approximation for the final estimates.

The conditional age distribution of SARI cases may nevertheless display multimodal patterns that vary across locations and epidemiological periods. A natural extension is to replace the lognormal working model with a flexible conditional density estimator, such as FlexCode [40], which estimates conditional densities through basis expansions whose coefficients are estimated by regression. This would allow the age distribution used by SViDAS to accommodate more complex shapes. Its impact on the accuracy of viral proportion estimates and on computational cost would require further evaluation.

### Scope and Interpretation of the Estimates

To ensure the accurate application of this framework, it is essential to consider the context in which these viral proportions are estimated. First, the understanding of viral distribution requires caution due to its sensitivity to testing strategies and resource availability. Comparing distributions across different localities presupposes a uniformity in testing policies, assuming that individuals with similar clinical conditions are tested equivalently in all compared areas. Similarly, contrasting proportions over time within the same locality assumes that testing protocols have remained stable. This implies a stability in the testing ratio for different viruses during the analyzed period, without shifts in the prioritization or exclusion of specific viral targets. Consequently, differential testing protocols may bias the observed viral detections, particularly when specific viral targets are prioritized or omitted (e.g., if hospitals test for SARS-CoV-2 but not RSV). Therefore, the estimates should be interpreted as the relative distribution among detected positive cases under the prevailing testing practices.

Second, the proportion of different viruses among SARI cases should be interpreted as the etiological profile of severe respiratory infections, rather than as a direct measure of viral circulation in the general population. This distinction arises because respiratory viruses differ in their transmission dynamics and in their probability of progressing to severe disease. A less virulent pathogen may be more prevalent in the general population than among hospitalized SARI cases, whereas a pathogen associated with more severe outcomes may be overrepresented among SARI notifications. This severe-disease profile is precisely the intended target for hospital surveillance and resource planning, but it should not be interpreted as a direct proxy for general community circulation.

### Computational Considerations

From an operational perspective, the computational efficiency was compatible with the intended deployment under limited resources. Using a standard notebook (Windows 11 Pro, AMD Ryzen 7 5700U, 16GB RAM), the models for the final epidemiological weeks of 2024—which comprised the largest dataset volume (2022–2024)—required less than two minutes to train. However, the retraining strategy requires access to the full historical dataset. As the SIVEP-Gripe database grows, the retraining time may eventually necessitate incremental learning strategies or more aggressive data subsampling.

### Practical Implications and Future Directions

The practical utility of this framework is demonstrated by its implementation in the PySRAG package and a dedicated web application. For public health authorities, the ability to query viral proportions for specific ages (e.g., ‘1 year’) supports targeted planning decisions, such as the timing of RSV prophylaxis or pediatric ICU allocation, which are often obscured in aggregate “all-ages” reports.

Future work should focus on integrating this viral proportion estimator with models for nowcasting total SARI case counts. By combining ViDAS estimates of viral proportions with nowcast estimates of SARI cases, it would be possible to obtain real-time estimates of virus-specific SARI burden. Additionally, incorporating meteorological data could further refine the spatial-temporal capture of seasonal respiratory viruses. Finally, extending the interpretability analysis using the embedding space could provide automated textual explanations for surveillance alerts, increasing trust and adoption among non-technical health officials.

Overall, the results indicate that ViDAS provides a complementary framework for timely and granular surveillance of the viral composition of SARI in Brazil. By combining spatial, temporal, and demographic information, the framework improves upon empirical aggregation, remains informative under reporting delays and local data scarcity, and provides mechanisms for interpreting individual predictions. These characteristics support its use for high-resolution surveillance of respiratory viruses among virus-positive SARI cases.

## Data Availability

All data produced in the present study are available upon reasonable request to the authors.

https://dadosabertos.saude.gov.br/dataset/srag-2019-a-2026

## 5. Declaration of generative AI and AI-assisted technologies in the manuscript preparation process

During the preparation of this work the authors used ChatGPT (OpenAI) and Gemini (Google) to assist with language editing and text clarity. After using this tool/service, the authors reviewed and edited the content as needed and take full responsibility for the content of the published article.

## Funding

This study was financed in part by the Coordenação de Aperfeiçoamento de Pessoal de Nível Superior – Brasil (CAPES) – Finance Code J.F.A. Silva received institutional support from Banco do Brasil through a program providing partial release from professional duties with continued remuneration during his doctoral studies. R.I. was supported by the Brazilian National Council for Scientific and Technological Development (CNPq) [grant numbers 422705/2021-7, and 305065/2023-8] and the São Paulo Research Foundation (FAPESP) [grant number 2023/07068-1 and 2025/24620-5]. The funding and supporting organizations had no role in the study design, data collection, analysis and interpretation of data, writing of the manuscript, or decision to submit the article for publication.

## Acknowledgments

This work was developed in part as part of the doctoral thesis of J.F.A. Silva, defended in 2026 in the Interinstitutional Graduate Program in Statistics (PIPGEs), jointly offered by UFSCar and USP.

## Notes

### Competing Interest Statement

The authors have declared no competing interest.

### Author Declarations

The study used ONLY openly available human data that were originally located at: https://dadosabertos.saude.gov.br/dataset/srag-2019-a-2026

