## Supplementary Material for "ViDAS: Timely and Granular Estimation of Respiratory Virus Distributions in SARI Surveillance in Brazil"

Monitoring Viral Infections in SARI Patients in Brazil

**Contents**

**S1 Model Configuration and Training Strategy** **2**

**S2 Geographic Coordinates and Tree-Based Models** **4**

**S3 Dataset Structure** **6**

**S4 Estimating Virus Distribution in Age Intervals: Derivation and Implementation Details** **6**

### S1. Model Configuration and Training Strategy

To construct the estimator, we utilized the Gradient Boosting Decision Tree (GBDT) classification algorithm via the LightGBM API [1]. This choice is based on the algorithm's ability to handle multidimensional data, model non-linearities, capture complex interactions among covariates, provide probabilistic classification, and deliver excellent computational performance [2].

The configuration of the LightGBM model involves defining several hyperparameters that are crucial for its performance. These settings are detailed below:

- `boosting_type="gbdt"`: Defines the type of boosting LightGBM uses. `"gbdt"` stands for "Gradient Boosted Decision Tree". This implies the algorithm is based on constructing decision trees to improve model performance. Trees are built in iterative stages, where each new tree is trained to correct errors made by previous ones.

- `objective="multiclass"`: Defines the model's objective function to be optimized during training. Here, `"multiclass"` indicates that LightGBM is configured to solve a multiclass classification problem, where data examples belong to more than two classes.

- `learning_rate=0.1`: Defines the algorithm's learning rate, i.e., how much the model learns from each added tree. In this case, the learning rate is set to 0.1.

- `n_estimators=5000`: Specifies the number of estimators (decision trees) to be created during training. The model is set to create up to 5000 trees.

- `max_depth=3`: Limits the maximum depth of decision trees. Setting `"max_depth"` to 3 prevents trees from growing beyond a depth of 3 levels, meaning each tree will have a maximum of $2^3$ leaves.

- `min_data_in_leaf=30`: Defines the minimum number of data points (samples) required in a leaf (terminal node). A value of 30 means a leaf will only be created if it contains at least 30 samples.

- `feature_fraction=0.8`: Controls the fraction of features (covariates) selected randomly in each iteration (tree construction). With a value of

0.8, LightGBM will randomly select 80% of the features before training each tree.

- `min_data_in_bin=10`: Specifies the minimum number of data points in each bin used to construct the histogram during training. Histograms are used in gradient boosting histogram algorithms like LightGBM to accelerate training. A value of 10 means a bin will only be created if it contains at least 10 data points.

The choice of `boosting_type="gbdt"` allows the model to effectively partition the variable space, particularly regarding the segmentation of epidemiological weeks, geographic coordinates, municipal population size, and patient age. This results in the creation of multiple local models or groups, each focusing on specific areas of the variable space, enabling the algorithm to handle different behaviors and patterns in the data.

The selection of `learning_rate=0.1` was strategic to ensure the efficiency of our solution within the monitoring dashboard, as reducing it affects both training and inference performance. During training, a lower `learning_rate` means each additional tree contributes less to correcting errors from previous trees, resulting in slower training as more iterations are needed to reach the optimal solution. Conversely, during the application of the trained model, a lower `learning_rate` may require a larger number of trees to obtain precise predictions, potentially increasing prediction time.

To evaluate model performance and determine the optimal number of boosting iterations, we adopted a holdout validation strategy to ensure the computational efficiency required for weekly updates, initially splitting the data into training (80%) and validation (20%) sets. To prevent overfitting, we employed early stopping as a criterion to halt the boosting iterations. Additionally, we incorporated the `multi_logloss` (multi-class logarithmic loss) evaluation metric as part of the stopping criterion; training is interrupted when the metric stops improving and begins to increase on the validation set. Once the optimal number of iterations is identified through this validation process, the final model is retrained using this number of iterations and the full dataset (100% of the data) to maximize the information available for estimation.

It is worth noting that this classifier algorithm with these configurations is available in the Python package PySRAG [3].

### S2. Geographic Coordinates and Tree-Based Models

The decision to employ geographic coordinates, such as latitude and longitude, instead of indicator variables (one-hot encoding) to represent location, is grounded in considerations related to the application of tree-based estimators. This section outlines the justification underlying this approach and highlights the associated advantages.

The primary reason for utilizing geographic coordinates is the preservation of distance relationships. Employing coordinates is fundamental to maintaining the spatial relationship between points. The use of indicator variables would result in the loss of information regarding the spatial proximity between observations. Consequently, there would be no differentiation between nearby or distant municipalities, potentially leading to the inadequate grouping of municipalities without considering their geographic proximity.

It is important to emphasize that this section focuses exclusively on raw latitude and longitude variables to demonstrate that, even without complex transformations, tree-based models can autonomously generate geographically coherent partitions. For instance, consider $c_{lat}$ as the number of partitions (splits) in the latitude coordinate and $c_{lon}$ as the number of partitions in the longitude coordinate. The combination of these partitions results in a total of $c_{lat} + c_{lon} + 1$ distinct regions (groups). This approach creates coherent groups that respect the distance between municipalities in the geographic space, as demonstrated in Figure S1.

While Figure S1 illustrates the rectangular partitions characteristic of individual decision trees, the application of Gradient Boosting Decision Trees (GBDT) fundamentally refines this spatial representation. Although a single tree produces discrete "jumps" between regions, the boosting process—by iteratively adding trees that correct previous errors—smooths these boundaries. As the number of boosting iterations increases and the learning rate decreases, the initially jagged curves separating regions become increasingly fine-grained, allowing for a smoother and nuanced modeling of spatial risk that approximates a continuous surface, rather than relying on the abrupt changes typical of single trees.

Another reason for this choice is computational efficiency. Using indicator variables to represent every municipality (in the case of Brazil, 5,570 municipalities) would lead to the creation of a massive number of features, many of which would be sparse. This would significantly slow down the es-

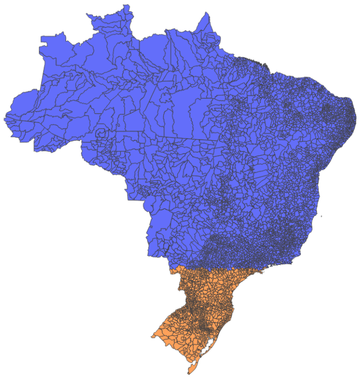

(a) 2 groups, $c_{lat} = 1$ and $c_{lon} = 0$.

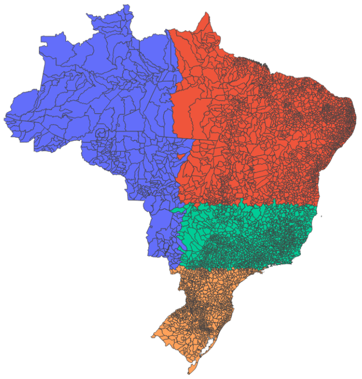

(b) 4 groups, $c_{lat} = 2$ and $c_{lon} = 1$.

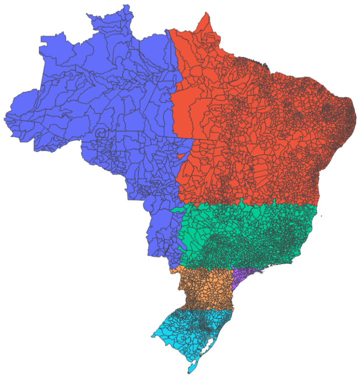

(c) 6 groups, $c_{lat} = 3$ and $c_{lon} = 2$

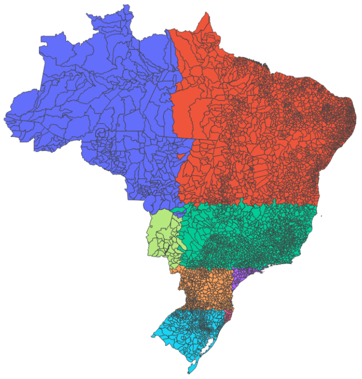

(d) 8 groups, $c_{lat} = 4$ and $c_{lon} = 3$

Figure S1: Example of the partitioning of the Brazilian geographic space based on municipality latitude and longitude coordinates. We observe that this partitioning generates groups consistent with the distances between municipalities.

timation process, depending on the algorithm used. Therefore, the use of geographic coordinates reduces the dimensionality of the data, making the process computationally more efficient.

In summary, the use of geographic coordinates and spatial partitioning offers an effective method for handling data scarcity in certain municipalities. Even when information is limited or absent for a specific municipality, it is possible to assign estimates based on information from neighboring municipalities within the same spatial partition.

### S3. Dataset Structure

The dataset $\mathbb{D}$ described in Section 2.2 of the main manuscript contains the epidemiological, demographic, and geographical covariates used by the proposed methodology. Table S1 provides an illustrative sample of its structure.

Table S1: Example illustrating the structure of dataset $\mathbb{D}$.

| Virus | Week | Patient's | City | | State | | Region | | Population |
|---|---|---|---|---|---|---|---|---|---|
| | 1st Sympt. | Age | Lat. | Long. | Lat. | Long. | Lat. | Long. | Size |
| FLUA | 1 | 68 | -23.87 | -47.99 | -23.54 | -46.63 | -21.64 | -43.53 | 32,062 |
| SARS2 | 3 | 56 | -23.54 | -46.63 | -23.54 | -46.63 | -21.64 | -43.53 | 12,200,180 |
| SARS2 | 4 | 72 | -10.91 | -37.07 | -10.91 | -37.07 | -7.31 | -37.99 | 605,309 |
| RSV | 5 | 0.17 | -16.82 | -49.24 | -16.67 | -49.25 | -17.12 | -51.98 | 500,760 |
| METAP | 12 | 2 | -23.54 | -46.63 | -23.54 | -46.63 | -21.64 | -43.53 | 12,200,180 |
| SARS2 | 26 | 46 | -23.59 | -48.05 | -23.54 | -46.63 | -21.64 | -43.53 | 166,959 |
| FLUA | 42 | 8 | -25.42 | -49.27 | -25.42 | -49.27 | -27.68 | -49.68 | 1,871,789 |
| Others | 46 | 68 | -14.67 | -40.47 | -12.97 | -38.51 | -7.31 | -37.99 | 23,290 |
| SARS2 | 47 | 43 | -30.03 | -51.22 | -30.03 | -51.22 | -27.68 | -49.68 | 1,404,269 |
| RSV | 48 | 0.83 | -23.54 | -46.63 | -23.54 | -46.63 | -21.64 | -43.53 | 12,200,180 |

### S4. Estimating Virus Distribution in Age Intervals: Derivation and Implementation Details

This section provides the complete methodological details for estimating virus distributions over patient age intervals, complementing Section 2.3.2 of the main manuscript. Specifically, it presents the derivation of the conditional virus distribution over an age interval, the estimation of the conditional age distribution through GBDT models, and the numerical procedure used to integrate the pointwise virus estimator over the corresponding age distribution.

#### *S4.1. Derivation of the Virus Distribution Conditioned on Age Intervals*

Our objective is to derive the expression for the conditional probability $\mathbb{P}_Y(y|X_j \in [a,b], \boldsymbol{X}_{-j} = \boldsymbol{x}_{-j})$, which represents the probability of observing virus $y$ given that the predictor variable $X_j$ (patient's age) falls within the interval $[a,b]$, holding constant the other covariates.

We define the notation where:

- $\boldsymbol{X} = (X_1, X_2, \ldots, X_p)$ is the vector of continuous predictor variables;

- $Y$ is the categorical random variable representing the virus type;
- $X_j$ is the specific variable of interest within the predictor set (patient's age);
- $\boldsymbol{X}_{-j} = (X_1, \ldots, X_{j-1}, X_{j+1}, \ldots, X_p)$ represents the vector of all predictor variables excluding $X_j$;
- $f_{X_j}(\cdot|\boldsymbol{x}_{-j})$ denotes the conditional probability density function of $X_j$ given the remaining covariates $\boldsymbol{x}_{-j}$;
- $F_{X_j}(\cdot|\boldsymbol{x}_{-j})$ denotes the corresponding conditional cumulative distribution function.

The derivation proceeds as follows:

$$\begin{aligned}
\mathbb{P}_Y(y|X_j \in [a,b], \boldsymbol{x}_{-j}) &= \int_{-\infty}^{\infty} \mathbb{P}_Y(y|X_j = x, \boldsymbol{x}_{-j}) f_{X_j}(x|X_j \in [a,b], \boldsymbol{x}_{-j})\, dx \\
&= \int_{-\infty}^{\infty} \mathbb{P}_Y(y|X_j = x, \boldsymbol{x}_{-j}) \frac{\mathbb{I}(x \in [a,b]) f_{X_j}(x|\boldsymbol{x}_{-j})}{\mathbb{P}(X_j \in [a,b]|\boldsymbol{x}_{-j})}\, dx \\
&= \frac{\int_{-\infty}^{\infty} \mathbb{P}_Y(y|X_j = x, \boldsymbol{x}_{-j}) \mathbb{I}(x \in [a,b]) f_{X_j}(x|\boldsymbol{x}_{-j})\, dx}{\mathbb{P}(X_j \in [a,b]|\boldsymbol{x}_{-j})} \\
&= \frac{\int_a^b \mathbb{P}_Y(y|X_j = x, \boldsymbol{x}_{-j}) f_{X_j}(x|\boldsymbol{x}_{-j})\, dx}{F_{X_j}(b|\boldsymbol{x}_{-j}) - F_{X_j}(a|\boldsymbol{x}_{-j})}.
\end{aligned}$$

*S4.2. Estimation of Conditional Density Parameters via GBDT*

The true conditional density $f_{X_j}(x_j|\boldsymbol{x}_{-j})$ is unknown and must be estimated. Alternative approaches for conditional density estimation include nonparametric methods such as FlexCode or quantile regression-based techniques [4, 5, 6]. While flexible, these methods typically require training a large ensemble of estimators—often necessitating dozens of distinct models to determine basis coefficients or specific quantile levels. Given our application's requirement for rapid response times and computational efficiency, we opted for a semiparametric approach. We simplify the estimation process by assuming a parametric distribution, which reduces the computational burden to estimating only the parameters of the distribution.

Since patient age is a strictly positive variable, we assume that it follows a lognormal distribution conditioned on location variables and the epidemiological week of symptom onset [7, 8]. That is,

$$X_j|\boldsymbol{x}_{-j} \sim LN\left(\mu(\boldsymbol{x}_{-j}), \sigma^2(\boldsymbol{x}_{-j})\right).$$

Consequently, its logarithm, $W = \log(X_j)$, is normally distributed [7]:

$$W|\boldsymbol{x}_{-j} \sim N\left(\mu(\boldsymbol{x}_{-j}), \sigma^2(\boldsymbol{x}_{-j})\right).$$

Under this assumption, the conditional density of patient age can be estimated by determining only the conditional mean and variance parameters [6]. We use an approach based on [9] to estimate these parameters. Consider the model:

$$W_i = \mu(\boldsymbol{x}_{-j,i}) + \sigma(\boldsymbol{x}_{-j,i})\epsilon_i,$$

where we define the auxiliary variables

$$Z_i = \log\left[(W_i - \mu(\boldsymbol{x}_{-j,i}))^2\right], \quad \delta_i = \log(\epsilon_i^2).$$

From these definitions, we have the relationship

$$Z_i = \log\left(\sigma^2(\boldsymbol{x}_{-j,i})\right) + \delta_i,$$

indicating that the estimate of $\log \sigma^2(\boldsymbol{x}_{-j})$ can be obtained by regressing the squared residuals (on a logarithmic scale) on $\boldsymbol{x}_{-j}$. Specifically, we proceed as follows:

1. Initially estimate the function $\mu(\boldsymbol{x}_{-j})$, representing the conditional mean estimate, using a GBDT estimator with $L_2$ loss for the objective function, obtaining $\hat{\mu}(\boldsymbol{x}_{-j})$.
2. Using this estimate, calculate the residuals and define:

$$Z_i = \log\left[(W_i - \hat{\mu}(\boldsymbol{x}_{-j,i}))^2\right].$$

3. Then regress the values $Z_i$ on the explanatory variables $\boldsymbol{x}_{-j,i}$, again using a GBDT estimator with $L_2$ loss for the objective function. This step yields an estimated function $\hat{q}(\boldsymbol{x}_{-j})$, corresponding to $\log \sigma^2(\boldsymbol{x}_{-j})$. Finally, the estimate of the conditional variance is given by

$$\hat{\sigma}^2(\boldsymbol{x}_{-j}) = e^{\hat{q}(\boldsymbol{x}_{-j})}.$$

Therefore, the estimated conditional density for the logarithm of patient age is given by

$$\hat{f}_W(w|\boldsymbol{x}_{-j}) = \frac{1}{\sqrt{2\pi\hat{\sigma}^2(\boldsymbol{x}_{-j})}} \exp\left(-\frac{(w - \hat{\mu}(\boldsymbol{x}_{-j}))^2}{2\hat{\sigma}^2(\boldsymbol{x}_{-j})}\right).$$

Similarly, the estimated conditional lognormal density for patient age is

$$\hat{f}_{X_j}(x_j|\boldsymbol{x}_{-j}) = \frac{1}{x\sqrt{2\pi\hat{\sigma}^2(\boldsymbol{x}_{-j})}} \exp\left(-\frac{(\log(x_j) - \hat{\mu}(\boldsymbol{x}_{-j}))^2}{2\hat{\sigma}^2(\boldsymbol{x}_{-j})}\right),$$

and the corresponding estimated cumulative distribution function is

$$\hat{F}_{X_j}(x_j|\boldsymbol{x}_{-j}) = \Phi\left(\frac{\log(x_j) - \hat{\mu}(\boldsymbol{x}_{-j})}{\hat{\sigma}(\boldsymbol{x}_{-j})}\right) \tag{S1}$$

where $\Phi(\cdot)$ is the cumulative distribution function of the standard normal distribution $N(0,1)$.

*S4.3. Numerical Integration Using the Probability-Weighted Midpoint Rule*

Having estimated the conditional distribution of viruses using the probabilistic estimator $\hat{g}(\boldsymbol{x})$ and the conditional density of patient age through the lognormal model, we integrate both estimators to obtain the virus distribution across an age interval. The corresponding estimator is

$$\hat{\mathbb{P}}_Y(y|X_j \in [a,b], \boldsymbol{x}_{-j}) = \frac{\int_a^b \hat{g}(x, \boldsymbol{x}_{-j})\hat{f}_{X_j}(x|\boldsymbol{x}_{-j})\,dx}{\hat{F}_{X_j}(b|\boldsymbol{x}_{-j}) - \hat{F}_{X_j}(a|\boldsymbol{x}_{-j})}. \tag{S2}$$

This estimator reflects the average predicted virus distribution over the age interval $[a,b]$, weighted by the likelihood of observing each age within that interval, conditional on the remaining covariates. The denominator in (S2) can be computed directly using the estimated cumulative distribution function in (S1). However, a numerical approximation is required to evaluate the integral in the numerator.

To estimate this integral, we subdivide the interval $[a,b]$ into $s-1$ subintervals using a sequence of ordered points $a = r_1 < r_2 < \cdots < r_s = b$. Then,

$$\int_a^b \hat{g}(x, \boldsymbol{x}_{-j})\hat{f}_{X_j}(x|\boldsymbol{x}_{-j})\,dx = \sum_{i=1}^{s-1}\int_{r_i}^{r_{i+1}} \hat{g}(x, \boldsymbol{x}_{-j})\hat{f}_{X_j}(x|\boldsymbol{x}_{-j})\,dx. \tag{S3}$$

Assuming that each subinterval has a sufficiently small length $h_i = r_{i+1} - r_i$, it is reasonable to approximate the function $\hat{g}(x, \boldsymbol{x}_{-j})$ within the subinterval $(r_i, r_{i+1})$ by evaluating it at a point $x^* \in [r_i, r_{i+1}]$, such as the midpoint $x^* = (r_i + r_{i+1})/2$. Thus,

$$\int_{r_i}^{r_{i+1}} \hat{g}(x, \boldsymbol{x}_{-j})\hat{f}_{X_j}(x|\boldsymbol{x}_{-j})\,dx \approx \hat{g}\left(\frac{r_i + r_{i+1}}{2}, \boldsymbol{x}_{-j}\right)\left[\hat{F}_{X_j}(r_{i+1}|\boldsymbol{x}_{-j}) - \hat{F}_{X_j}(r_i|\boldsymbol{x}_{-j})\right].$$

Therefore, the integral in (S3) can be approximated by

$$\int_a^b \hat{g}(x, \boldsymbol{x}_{-j}) \hat{f}_{X_j}(x|\boldsymbol{x}_{-j})\, dx \approx \sum_{i=1}^{s-1} \hat{g}\left(\frac{r_i + r_{i+1}}{2}, \boldsymbol{x}_{-j}\right) \left[\hat{F}_{X_j}(r_{i+1}|\boldsymbol{x}_{-j}) - \hat{F}_{X_j}(r_i|\boldsymbol{x}_{-j})\right]. \tag{S4}$$

Consequently,

$$\hat{\mathbb{P}}_Y(y|X_j \in [a,b], \boldsymbol{x}_{-j}) \approx \frac{\sum_{i=1}^{s-1} \hat{g}\left(\frac{r_i+r_{i+1}}{2}, \boldsymbol{x}_{-j}\right) \left[\hat{F}_{X_j}(r_{i+1}|\boldsymbol{x}_{-j}) - \hat{F}_{X_j}(r_i|\boldsymbol{x}_{-j})\right]}{\hat{F}_{X_j}(b|\boldsymbol{x}_{-j}) - \hat{F}_{X_j}(a|\boldsymbol{x}_{-j})}.$$

This approximation is analogous to applying the *midpoint rule* [10] to a Riemann–Stieltjes integral. The key difference from the classical formulation is that, rather than using the subinterval widths $r_{i+1} - r_i$, we use the probability mass of each region $[r_i, r_{i+1}]$ under the estimated cumulative distribution function $\hat{F}_{X_j}(\cdot|\boldsymbol{x}_{-j})$ as the integrator, while evaluating $\hat{g}(\cdot, \boldsymbol{x}_{-j})$ at the midpoint. In contrast, the classical midpoint rule over a Riemann integral would evaluate the product $\hat{g}(\cdot, \boldsymbol{x}_{-j})\hat{f}_{X_j}(\cdot|\boldsymbol{x}_{-j})$ and multiply it by the corresponding subinterval width.

### S5. Definition of the Brier Score

The Brier score is a proper scoring rule used to assess the quality and calibration of probabilistic predictions [11]. For a multi-class classification problem, it is defined as the mean squared difference between the predicted probabilities and the actual outcomes. Given a sample of size $n$, the Brier score is calculated as follows [6]:

$$\text{Brier Score} = \frac{1}{n} \sum_{i=1}^{n} \sum_{k=1}^{K} \left(\hat{g}_k(\boldsymbol{x}_i) - \mathbb{I}(y_i = k)\right)^2$$

where

- $n$ is the total number of observations.
- $K$ is the total number of classes (viruses).
- $\boldsymbol{x}_i$ is the feature vector for observation $i$.
- $y_i$ is the true class for observation $i$.

- $\hat{g}_k(\boldsymbol{x}_i)$ is the predicted probability that observation $i$ belongs to class $k$.

- $\mathbb{I}(y_i = k)$ is the indicator function, which is 1 if the true class of observation $i$ is $k$, and 0 otherwise.

A lower Brier score indicates better estimator performance, with a score of 0 representing a perfect fit in terms of both prediction and calibration.

### S6. Supplementary Results

#### *S6.1. Observed Virus Distribution*

The distribution of respiratory viruses observed during the 2022–2024 study period is summarized in Table S2. The proportions were calculated using the consolidated SIVEP-Gripe data released by the Brazilian Ministry of Health on May 5, 2025, which were used as the reference dataset throughout the analyses presented in the main manuscript.

Table S2: Observed virus proportion for the period from 2022 to 2024, based on SIVEP-Gripe data released by the Brazilian Ministry of Health on May 5, 2025.

| **Virus** | **Proportion** |
|---|---:|
| SARS-CoV-2 | 0.4469 |
| RSV | 0.2212 |
| Rhinovirus | 0.1388 |
| Influenza A | 0.0868 |
| Adenovirus | 0.0276 |
| Metapneumovirus | 0.0237 |
| Other Viruses | 0.0160 |
| Influenza B | 0.0158 |
| Parainfluenza 3 | 0.0109 |
| Bocavirus | 0.0083 |
| Parainfluenza 1 | 0.0018 |
| Parainfluenza 4 | 0.0013 |
| Parainfluenza 2 | 0.0010 |

*S6.2. Performance Evaluation of the Proposed Methodology*

This supplementary section provides additional methodological details and complete age-stratified results for the performance evaluation summarized in the main manuscript. The evaluation focuses on ANViDAS and SViDAS, which are capable of generating estimates for age intervals, a functionality not supported by NViDAS. Their performance was investigated through two complementary approaches: a graphical case study and a systematic quantitative evaluation of estimation errors.

*Evaluation Methodology*

The evaluation was designed to compare the performance of the proposed models (ANViDAS and SViDAS) and a Naive Model, using the Consolidated Data as a reference.

The **Consolidated Data**, representing the most accurate available epidemiological information, was constructed using the most complete and recent database released on May 5, 2025. For the graphical analysis, 95% confidence intervals for the weekly proportions were calculated using the Clopper–Pearson method, an exact binomial approach for proportions [12].

The graphical analysis focused on the municipality of São Paulo during the first 26 epidemiological weeks of 2022. The choice of a metropolis such as São Paulo was strategic, as the volume of data allows for the construction of robust Consolidated Data for each analyzed stratum. Estimates from ANViDAS, SViDAS, and the Naive Model were generated using data available on July 4, 2022 (epidemiological week 27), simulating a nowcasting scenario. The analysis was segmented by age ranges (overall, <2, 2–5, 5–15, 15–50, 50–65, and 65+ years) for SARS-CoV-2 and RSV, selected due to their high incidence.

The quantitative evaluation complements the graphical analysis by simulating the operational use of the models. The models were retrained with each new data release between January 1, 2022, and May 5, 2025, and their estimates were evaluated for epidemiological weeks within the 2022–2024 period. For each data release, a new set of estimators was generated and used to predict the 14-week window preceding its creation date, focusing the evaluation on the period in which data are most incomplete and subject to updates. Performance was measured using the Brier Score [11], a proper scoring rule that measures the quality of probabilistic predictions by calculating the mean squared difference between predicted probabilities and observed outcomes (1 for positive and 0 for negative) [6]. Scores range from 0 to 1, with lower values indicating better probabilistic predictions. Estimates were compared against individual outcomes from the Consolidated Data. To ensure comparability, imputation strategies were adopted for cases of missing data: for ANViDAS, the midpoint of the age range was used to generate the estimate; for the Naive Model, in the rare event of a complete absence of data for a State and epidemiological week, a discrete uniform distribution was assumed for the virus distribution.

*Graphical Analysis: Viral Dynamics and Model Performance*

Figure S2 and Figure S3, which present the complete age-stratified viral proportions for SARS-CoV-2 and RSV, reveal markedly distinct temporal and demographic dynamics. A clear negative correlation is observed between the proportions of the two viruses, where an increase in one frequently coincides with a decrease in the other. This dynamic is a consequence of the proportional data structure and the predominant incidence of these two agents during the period, reflecting their circulation in different subpopulations and time frames rather than direct viral competition.

ANViDAS and SViDAS demonstrate consistently superior graphical performance compared with the Naive Model, with estimates that closely track the Consolidated Data curves and often fall within their confidence intervals. SViDAS, by modeling age continuously, generates notably smooth curves, particularly in scenarios with fewer observations, where the Consolidated Data exhibit greater volatility and wider confidence intervals. ANViDAS, based on the empirical age distribution, also produces regular estimates, although it more directly reflects weekly fluctuations and cannot generate predictions in the absence of observations—a limitation illustrated, for example, in week 12 for the 50–65 age range in Figure S2 and Figure S3.

The Naive Model shows systematic and age-dependent errors. The visual analysis indicates that it overestimates the proportion of RSV in adults and the elderly and underestimates it in the pediatric population, while the opposite pattern occurs for SARS-CoV-2. Age stratification also reveals important epidemiological patterns, including the predominance of RSV in the pediatric population and the persistence of SARS-CoV-2 as the virus with the highest proportion among the elderly, even during periods of low overall circulation.

*Quantitative Analysis: Validation of Estimation Errors*

The quantitative evaluation corroborates and extends the findings of the

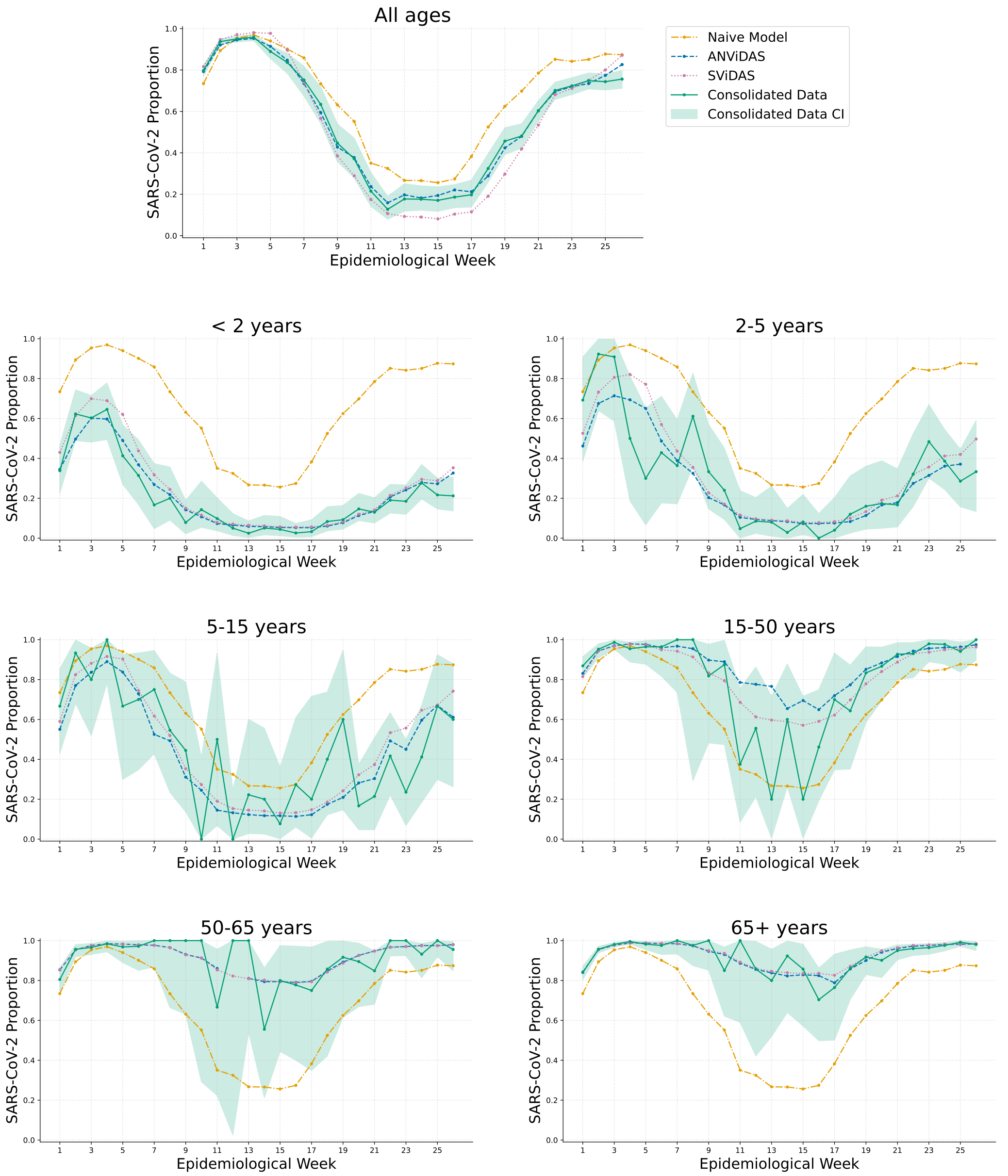

Figure S2: Estimated weekly proportion of SARS-CoV-2 among virus-positive SARI cases in São Paulo for the first 26 epidemiological weeks of 2022, stratified by age group. The figure compares the estimates from the proposed models (**ANViDAS** and **SViDAS**) and the Naive Model with the observed proportions from the Consolidated Data. Shaded areas represent the 95% confidence interval for the Consolidated Data. Both **ANViDAS** and **SViDAS** estimate the Consolidated Data more accurately than the Naive Model.

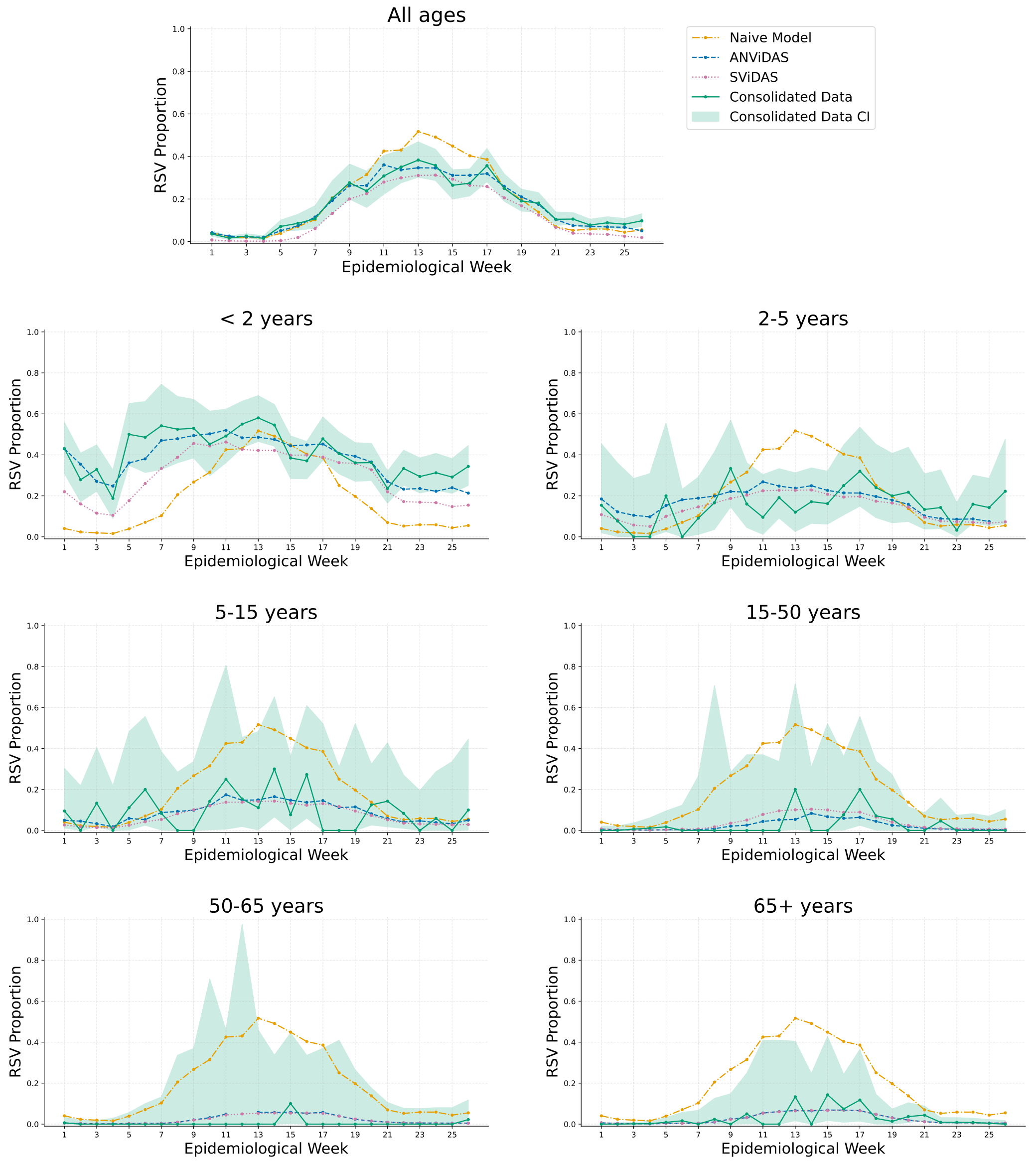

Figure S3: Estimated weekly proportion of RSV among virus-positive SARI cases in São Paulo for the first 26 epidemiological weeks of 2022, stratified by age group. The figure compares the estimates from the proposed models (**ANViDAS** and **SViDAS**) and the Naive Model with the observed proportions from the Consolidated Data. Shaded areas represent the 95% confidence interval for the Consolidated Data. Both **ANViDAS** and **SViDAS** estimate the Consolidated Data more accurately than the Naive Model.

graphical analysis. As shown in the main manuscript, ANViDAS and SViDAS outperform the Naive Model across all 13 viruses, with substantial error reductions for the most incident agents. For SARS-CoV-2, for example, the average Brier Score is reduced by more than 40% compared with the Naive Model. ANViDAS and SViDAS show very similar performance, with ANViDAS having a marginal advantage in most cases, indicating that both approaches to incorporating age distribution—empirical and modeled, respectively—are effective. The lower standard deviations observed for the most prevalent viruses also indicate greater stability in predictive performance across evaluations.

Table S3 details the source of these improvements by stratifying the error by age range. The analysis of RSV clearly illustrates the inadequacy of the Naive Model, which shows a high error (0.2549) in the $<$2-year age group, whereas the proposed models substantially reduce it (to approximately 0.18). Even among adults and the elderly, where RSV is less frequent, the proposed models reduce the Naive Model's error from approximately 0.08 to approximately 0.03.

For SARS-CoV-2, the age-stratified results quantify the systematic error of the Naive Model in the elderly population (65+ years), where it reaches its highest value (0.1897), while the proposed models reduce the error to approximately 0.102. For Rhinovirus and Influenza A, the proposed models also outperform the Naive Model across all age ranges, with the largest error for Rhinovirus occurring between 2 and 15 years, a pattern more accurately captured by ANViDAS and SViDAS. Together, these results quantify the systematic age-dependent errors of the Naive Model and further support the importance of age stratification.

Overall, the combined evaluation supports the ANViDAS and SViDAS methodologies, demonstrating their ability to reveal and quantify viral dynamics that are obscured by non-stratified approaches. The identification of specific demographic patterns, such as the predominance of RSV among children and the higher proportion of SARS-CoV-2 among older individuals, illustrates the epidemiological information gained by explicitly incorporating age stratification.

*S6.3. Analysis of the Effect of Delays on Estimates*

This supplementary section provides additional methodological details and complete results for the analysis of reporting delays summarized in the

Table S3: Mean Brier Score (and standard deviation) for the four most incident viruses, stratified by age range. The lowest error for each scenario is highlighted in bold.

| Age Range | Virus | Naive Model<br>Mean (SD) | ANViDAS<br>Mean (SD) | SViDAS<br>Mean (SD) |
|---|---|---|---|---|
| < 2 years | Influenza A | 0.0412 (0.0251) | **0.0296** (0.0149) | 0.0301 (0.0152) |
| | Rhinovirus | 0.1231 (0.0463) | **0.1164** (0.0434) | 0.1174 (0.0437) |
| | SARS-CoV-2 | 0.1722 (0.1072) | **0.0802** (0.0380) | 0.0814 (0.0389) |
| | RSV | 0.2549 (0.0561) | **0.1821** (0.0256) | 0.1873 (0.0264) |
| 2-5 years | Influenza A | 0.0690 (0.0400) | **0.0625** (0.0359) | 0.0626 (0.0358) |
| | Rhinovirus | 0.1985 (0.0388) | **0.1685** (0.0299) | 0.1694 (0.0299) |
| | SARS-CoV-2 | 0.1627 (0.0982) | **0.0782** (0.0411) | 0.0789 (0.0419) |
| | RSV | 0.1642 (0.0579) | **0.1454** (0.0505) | 0.1475 (0.0510) |
| 5-15 years | Influenza A | 0.1199 (0.0613) | **0.1032** (0.0523) | 0.1040 (0.0522) |
| | Rhinovirus | 0.2397 (0.0399) | **0.1747** (0.0255) | 0.1759 (0.0252) |
| | SARS-CoV-2 | 0.1579 (0.0797) | **0.0958** (0.0445) | 0.0980 (0.0466) |
| | RSV | 0.1102 (0.0612) | **0.0759** (0.0372) | 0.0769 (0.0376) |
| 15-50 years | Influenza A | 0.1371 (0.0992) | **0.1059** (0.0629) | 0.1067 (0.0632) |
| | Rhinovirus | 0.0812 (0.0513) | **0.0720** (0.0451) | 0.0728 (0.0458) |
| | SARS-CoV-2 | 0.1666 (0.0508) | **0.1269** (0.0405) | 0.1290 (0.0415) |
| | RSV | 0.0846 (0.0631) | **0.0295** (0.0235) | 0.0296 (0.0237) |
| 50-65 years | Influenza A | 0.1354 (0.1111) | **0.1009** (0.0699) | 0.1012 (0.0702) |
| | Rhinovirus | 0.0586 (0.0442) | **0.0473** (0.0360) | 0.0475 (0.0362) |
| | SARS-CoV-2 | 0.1725 (0.0665) | **0.1160** (0.0514) | 0.1163 (0.0517) |
| | RSV | 0.0837 (0.0655) | **0.0322** (0.0263) | 0.0323 (0.0264) |
| 65+ years | Influenza A | 0.1102 (0.1032) | **0.0848** (0.0684) | 0.0857 (0.0691) |
| | Rhinovirus | 0.0501 (0.0418) | **0.0358** (0.0318) | 0.0363 (0.0321) |
| | SARS-CoV-2 | 0.1897 (0.0871) | **0.1015** (0.0562) | 0.1029 (0.0574) |
| | RSV | 0.0819 (0.0659) | **0.0301** (0.0288) | 0.0303 (0.0290) |

main manuscript. We evaluated the impact of reporting delays on the performance of the estimates generated by the proposed methodology. To this end, we conducted an experiment simulating the operational use of NViDAS over time, mimicking a nowcasting setting in which predictions are initially generated from partial information and subsequently updated as data are consolidated. The analysis was based on weekly data releases from January 2022 to May 2025. For each release, a new NViDAS estimator was trained, reflecting the information available at that specific point in time. Performance for each epidemiological week was evaluated as a function of the reporting lag, defined as the difference, in weeks, between the data release date and the epidemiological week for which the estimate was generated.

To ensure a robust analysis of the most critical period, during which data are incomplete and subject to updates, the analysis focused on a retrospective window of up to 14 weeks of lag. Although lag 0 represents the current epidemiological week in the conceptual diagram, it was not included in the analysis because weekly data releases generally occur before that epidemiological week is complete. Therefore, the analysis started at lag 1. Only epidemiological weeks with estimates available for at least seven of the possible lags, including lag 1, were retained. Performance was measured using the Brier Score, comparing the predicted probabilities against the individual outcomes from the consolidated data released on May 5, 2025. To facilitate comparison across different epidemiological weeks, the Brier Scores for each virus were standardized using Min–Max scaling within each week. The conceptual design of this evaluation is illustrated in Figure S4.

The results, presented in Figure S5, show the average behavior of the standardized Brier Scores across reporting lags for the four most incident viruses: SARS-CoV-2, RSV, Rhinovirus, and Influenza A. A consistent pattern was observed across Brazil and its five macro-regions: estimation error decreased sharply during the first few weeks and subsequently tended to stabilize. This rapid improvement may partly reflect the increasing volume of available data, but may also be influenced by the dynamics of laboratory confirmation, whereby cases with higher clinical probability may be confirmed earlier and additional results are progressively incorporated into the surveillance system. As the data are consolidated, the estimates become increasingly precise.

The line graphs in Figure S5 indicate that, for the main viruses, the estimation error tends to stabilize after a lag of approximately 3 to 5 weeks. Table S4 quantifies this stabilization point, identifying, for each virus and

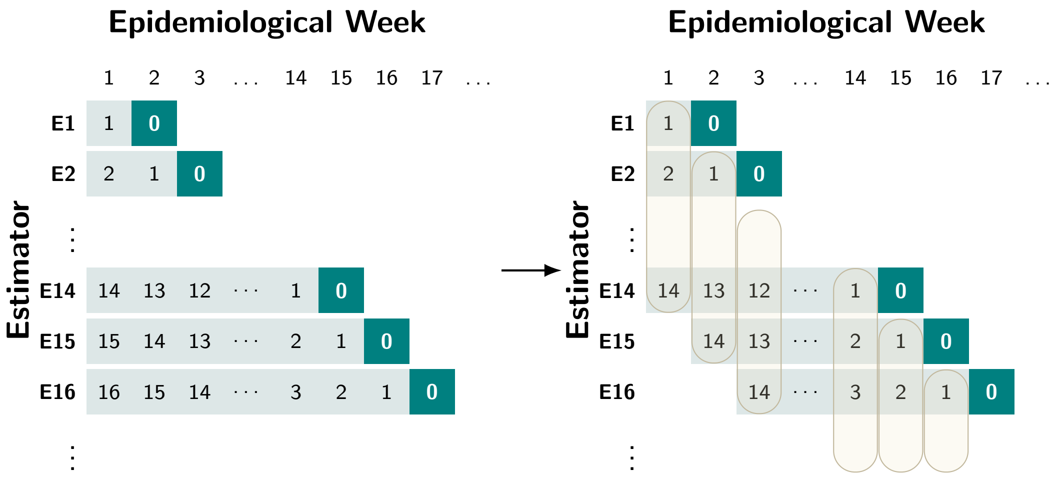

Figure S4: Conceptual diagram of the delay analysis methodology. The diagram on the left illustrates the estimation process, where each row (E1, E2,...) represents an estimator trained on a new weekly data release. The columns are epidemiological weeks, and the numbers within the cells indicate the lag (in weeks) of each estimate, with lag 0 highlighted as the week of the data release. The diagram on the right represents the final dataset used for the analysis, which is obtained after applying the following steps: (1) estimates with a lag greater than 14 weeks are filtered out; (2) only epidemiological weeks with at least seven estimates, including the one at lag 1, are retained; and (3) the resulting Brier scores are standardized for each epidemiological week.

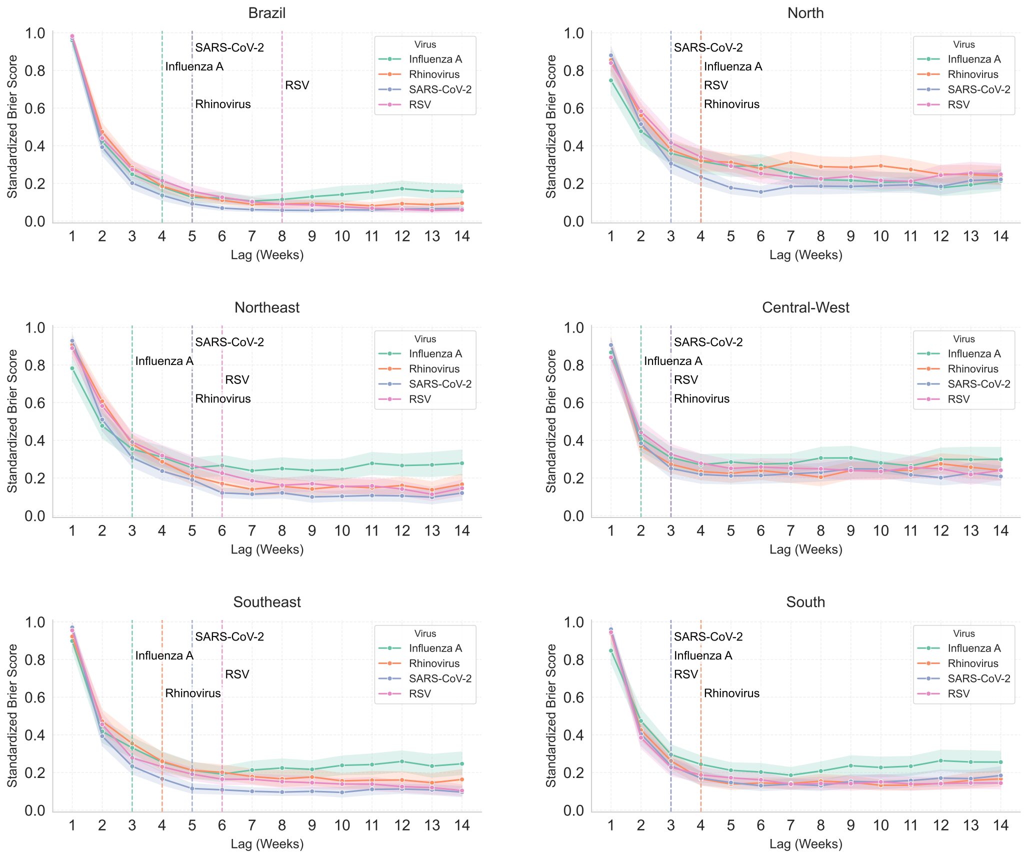

Figure S5: Impact of data reporting lag on the performance of virus estimates, measured by the Standardized Brier Score. Each panel shows the mean score (solid line) and its 95% confidence interval (shaded area) as a function of the lag in weeks for the four most incident viruses. The analysis is presented for Brazil as a whole and for its five macro-regions. The vertical dashed lines mark the stabilization point for each virus, corresponding to the values in Table S4. A consistent pattern is observed across all regions, with a sharp improvement in performance during the first few weeks, followed by progressive stabilization, with the stabilization point varying by virus and geographical region.

region, the smallest lag at which the bootstrapped 95% confidence interval [13] of the error estimate overlaps with that of the 14-week lag, suggesting that the error has reached a stable level. For Brazil as a whole, for example, estimates for SARS-CoV-2 and Rhinovirus stabilize at a lag of 5 weeks, while Influenza A stabilizes earlier, at 4 weeks.

Table S4: Minimum Stabilization Lag for Main Viruses by Region. The value represents the minimum lag at which the estimate's performance is considered stable. Stability is defined as the first point where the 95% confidence interval of the standardized Brier score for a given lag overlaps with the 95% CI of the 14-week lag.

| **Region** | **Virus** | | | |
|---|---|---|---|---|
| | **SARS-CoV-2** | **RSV** | **Rhinovirus** | **Influenza A** |
| Brazil | 5 | 8 | 5 | 4 |
| North | 3 | 4 | 4 | 4 |
| Northeast | 5 | 6 | 5 | 3 |
| Central-West | 3 | 3 | 3 | 2 |
| Southeast | 5 | 6 | 4 | 3 |
| South | 3 | 3 | 4 | 3 |

Furthermore, the mean standardized score generally reaches values between 0.2 and 0.4, on a scale from 0 to 1, from lag 3 onward in nearly all cases, while the minimum stabilized mean error typically lies between 0.1 and 0.3. Thus, by approximately the third week, the estimation error is already close to the lowest levels observed after further data consolidation in most cases. This pattern indicates that a substantial proportion of the improvement in estimation performance occurs early in the reporting process, while subsequent updates tend to yield comparatively smaller gains.

*S6.4. Interpretation and Explanation of the NViDAS Model*

This supplementary subsection provides the complete interpretation analysis of the NViDAS model, including the importance of predictor variables and the influence of key features, particularly age and epidemiological week, on the predicted outcome. The analysis was conducted using the most recent estimator, trained on the dataset released in May 2025.

*Analysis of Predictor Variable Importance*

First, we evaluated the global importance of each predictor variable by quantifying its total gain, which represents the overall improvement in prediction accuracy contributed by that feature across all the branches where it is used in a tree-based model such as GBDT [13]. The analysis reveals a clear hierarchy among the variables, as shown in Figure S6. Patient's Age emerges as the most influential predictor by a substantial margin, followed by the Epidemiological Week of the first symptoms. Geographical variables exhibit a tiered importance corresponding to their granularity, with city-level coordinates being more influential than state-level, which in turn are more important than region-level coordinates. This pattern indicates that the model effectively captures localized viral transmission dynamics. The Population Size of the notifying municipality also ranks as a relevant predictor, indicating an additional contribution of municipality size to the model.

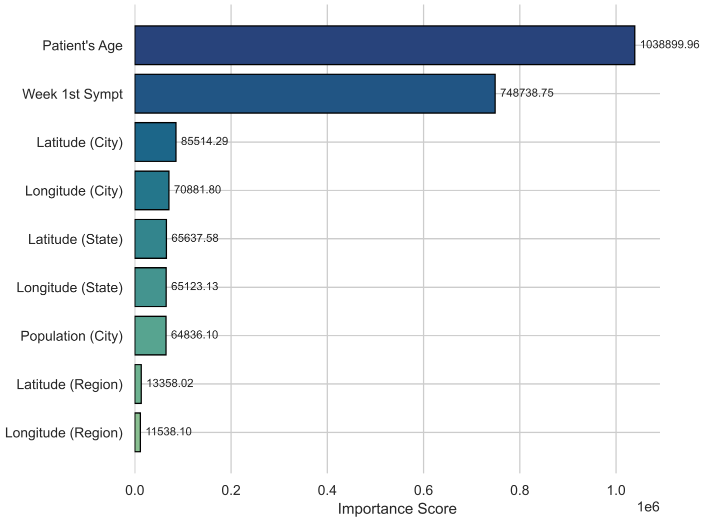

Figure S6: Predictor Variable Importance based on Total Gain. The chart illustrates the relative importance of each feature, with Patient's Age and Epidemiological Week emerging as the most influential predictors.

*Visualizing Predictor Effects with PDP and ICE Plots*

To visualize how the most influential variables affect the model's outputs, we generated Partial Dependence Plots (PDPs) and Individual Conditional Expectation (ICE) plots [14, 15], using data not included in the model training. The PDP illustrates the average effect of a feature on the predicted outcome, computed using a random sample of 1000 observations. To reveal the heterogeneity of this effect, ICE plots show the prediction for individual instances; for visual clarity, these plots were generated for a subsample of 100 observations.

Figure S7 presents these visualizations for the four main viruses, revealing distinct and clinically recognized epidemiological profiles.

- **SARS-CoV-2:** The model learned a strong positive correlation between age and the probability of SARS-CoV-2. The predicted likelihood is low for infants and increases steadily with age, reaching its peak in the elderly population (65+ years). This age-dependent risk profile is further corroborated by the analysis of temporal trends. During periods of high viral circulation, the epidemiological week plot consistently shows that the probability curves for older age groups (light green) are positioned at the top, confirming their heightened susceptibility.

- **Respiratory Syncytial Virus (RSV):** In stark contrast to SARS-CoV-2, the model identified a powerful negative correlation between age and RSV probability. The likelihood of infection is exceptionally high for neonates and infants—a vulnerability that drops sharply within the first few years of life before stabilizing at a low baseline for adolescents and adults. This finding aligns with established RSV epidemiology, where primary infections are most severe in the immunologically naive pediatric population. The epidemiological week plot reinforces this age stratification, showing that pediatric age groups (dark purple and blue) consistently dominate the seasonal peaks of RSV activity.

- **Rhinovirus:** The model captured a more complex, non-linear relationship for Rhinovirus, with the probability peaking in early-to-mid childhood (approximately 5-15 years) before gradually declining through adulthood. This pattern likely reflects the combined effects of intense social mixing in school environments and the gradual acquisition of immunity to numerous Rhinovirus serotypes over a lifetime. The corresponding epidemiological week plot supports this observation, showing a clear stratification where the probability curves for pediatric and adolescent groups remain generally higher than those for adults across different seasons.

- **Influenza A:** While the predicted probability of Influenza A shows a slight increase during childhood before leveling off, the time-series plot reveals an extremely strong seasonal signal, with sharp, distinct

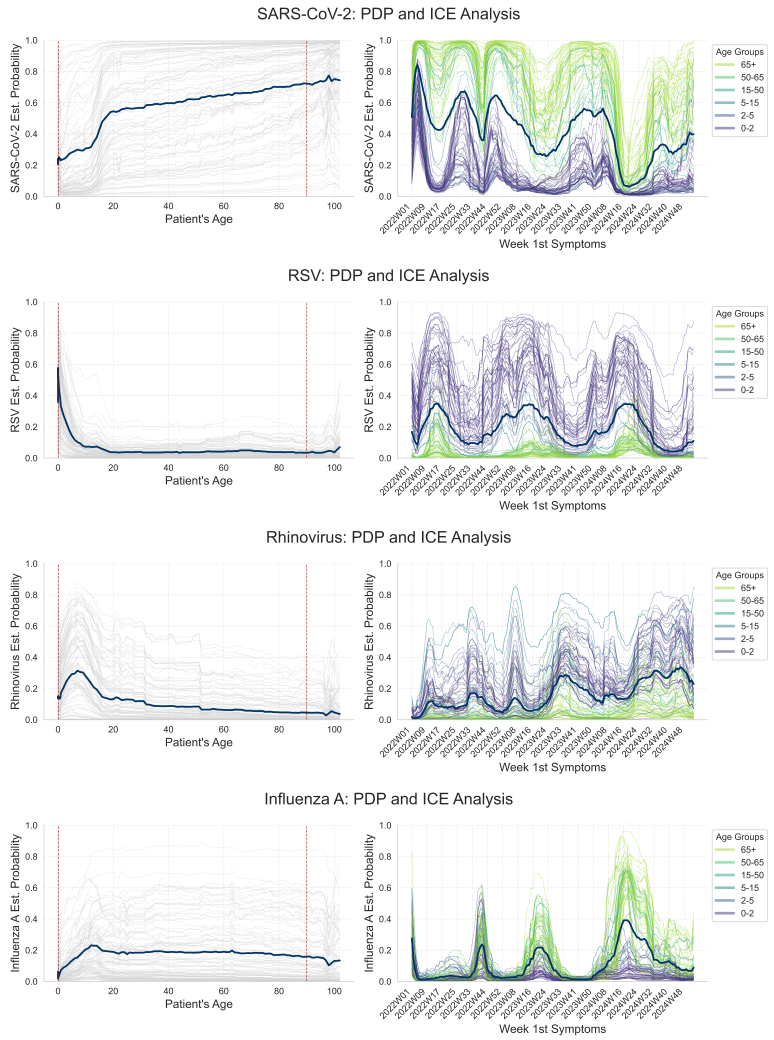

Figure S7: Learned effects of patient age and epidemiological week on the estimated probabilities for the four main viruses. For each virus, the left panel displays the effect of age, with the bold line representing the Partial Dependence Plot (PDP), faint lines showing Individual Conditional Expectation (ICE) plots, and red dashed vertical lines indicating the 5th and 95th percentiles of observed patient age, which delimit the central range of the empirical age distribution and highlight age regions with fewer observations. The right panel displays the effect of the epidemiological week, where the bold line is the PDP, and the colored ICE plots illustrate the interaction with different age groups as detailed in the legend. The plots reveal that the model has successfully learned distinct and clinically recognized epidemiological profiles for each virus.

peaks across all age groups. This indicates that for Influenza A, the epidemiological week is a more dominant predictor than the patient's age.

These visualizations confirm that the model has not simply identified age and time as important predictors, but has learned how their interaction shapes the probability of each virus, capturing distinct epidemiological signatures. In practice, these patterns corroborate the empirical observation that age-stratified weekly SARI trends can provide clues about the predominant virus: increases concentrated among infants are more compatible with RSV, increases among children and adolescents with Rhinovirus, increases concentrated among the elderly with SARS-CoV-2, and increases observed across most age groups with Influenza A.

*Local Interpretation in a Zero-Data Scenario*

To demonstrate the model's decision-making process in a data-scarcity scenario, we analyzed its predictions for hypothetical cases in Araporã, a municipality with a small population (approx. 8,500) located in the Triângulo Mineiro region of Minas Gerais, for which no SARI cases were recorded in 2024 after applying the study's filters. We investigated two distinct profiles for epidemiological week 22 of 2024.

**Case 1: Influenza A in a 30-Year-Old Adult.** For a hypothetical 30-year-old patient, the model assigned a high probability of 78.0% to Influenza A. To explain this, we applied the nearest-neighbor analysis in the model's embedding space, as described in Section 2.2.4. The 10 most similar training instances were from four nearby municipalities within the same state (Minas Gerais). Within this group, the observed proportion of Influenza A was 70.0%. A rule-based filter, defined by the minimum and maximum observed values for the neighbors' covariates (age: 15-83 years, population: 21k-102k, epidemiological weeks: 2024W22-2024W30, and geographic location in the Triângulo Mineiro region; see Figure S8), identified a slightly larger group of 13 cases, where the observed proportion of Influenza A was 76.9%, closely matching the model's prediction.

**Case 2: Respiratory Syncytial Virus (RSV) in a Neonate.** For a hypothetical neonate (age 0.01 years), the model predicted a 54.1% probability of RSV. The nearest-neighbor analysis identified a different set of influential cases: 10 infants from six other cities in Minas Gerais, where the observed RSV proportion was 60.0%. The corresponding rule-based filter,

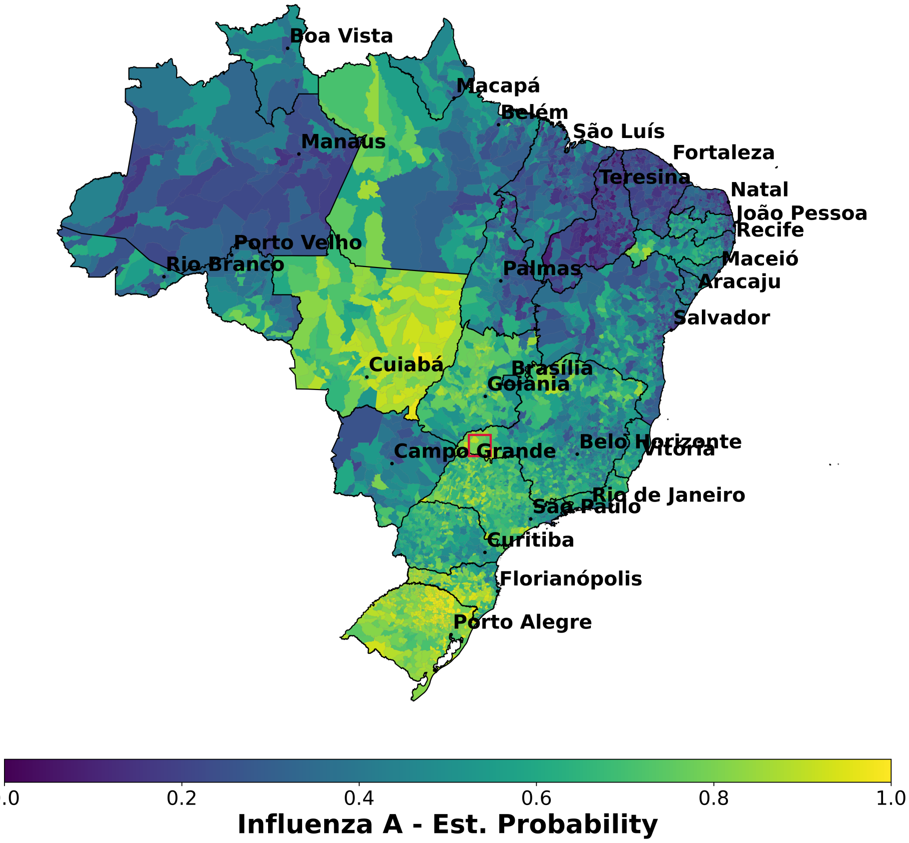

Figure S8: Spatial Individual Conditional Expectation (ICE) plot showing the predicted probability of Influenza A across geographic coordinates for a hypothetical 30-year-old patient. The overlaid rectangle delineates the spatial boundaries derived from the min-max rule applied to the nearest neighbors in the model's embedding space. The model anchors its high-probability prediction for this zero-data municipality by identifying a highly localized spatial cluster of similar adult cases within the Triângulo Mineiro region.

derived from the neighbors' characteristics (age: approx. 1 month, population: 81k-713k, epidemiological weeks: 2024W22-2024W23, and a geographic location in a wider region containing the Triângulo Mineiro and other areas in central and southern Minas Gerais, see Figure S9), identified a larger set of 23 infants, in which the observed RSV proportion was 52.2%, again aligning closely with the model's prediction.

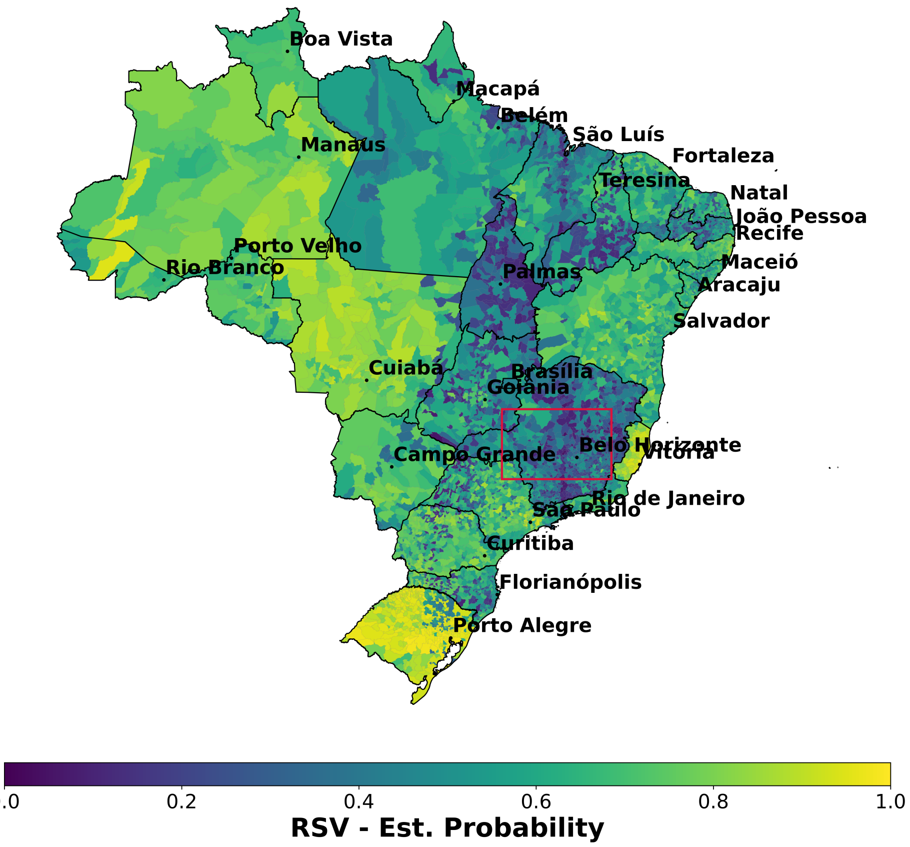

Figure S9: Spatial Individual Conditional Expectation (ICE) plot illustrating the predicted probability of Respiratory Syncytial Virus (RSV) across geographic coordinates for a hypothetical neonate (0.01 years old). The bounding rectangle represents the spatial filter defined by the most influential nearest neighbors. In contrast to the highly localized adult profile in Case 1, the model draws upon a broader regional area across central and southern Minas Gerais. Notably, rather than showing a dense concentration of high-probability hotspots within this bounding box, the plot reveals a more diffuse and generalized distribution of risk.

This case study reveals that in the absence of direct evidence, the model bases its predictions on data from demographically, temporally, and geographically similar profiles. The model inferred that the regional trend for an adult pointed to Influenza A, while for a neonate the dominant risk was RSV, with both predictions being consistent with the patterns observed among demographically, temporally, and geographically similar profiles.

**List of Notation**

| Notation | Description |
|---|---|
| *Data and Variables* | |
| $\mathbb{D}$ | Training dataset, defined as $\{(\boldsymbol{x}_i, y_i)\}_{i=1}^n$ |
| $\boldsymbol{X}, \boldsymbol{x}$ | Vector of predictor variables (random variable and realization, respectively) |
| $Y, y$ | Categorical target variable representing the virus type (random variable and realization, respectively) |
| $X_j$ | Specific predictor variable of interest (e.g., patient age) |
| $\boldsymbol{X}_{-j}, \boldsymbol{x}_{-j}$ | Vector of all predictor variables excluding $X_j$ (random variable and realization, respectively) |
| $\mathbb{I}(\cdot)$ | Indicator function, equal to 1 if the condition is true, 0 otherwise |
| *Probabilistic Estimators* | |
| $\mathbb{P}_Y(y\|\boldsymbol{x})$ | Conditional probability of observing virus $y$ given covariates $\boldsymbol{x}$ |
| $\hat{g}(\boldsymbol{x})$ | Multi-class probabilistic estimator (GBDT model) |
| $\hat{g}_k(\boldsymbol{x})$ | Estimated probability for virus class $k$ given $\boldsymbol{x}$ |
| $h_k(\boldsymbol{x})$ | Raw score (logit) for class $k$ produced by the ensemble |
| $b_{mk}(\boldsymbol{x})$ | Output of the $m$-th decision tree for class $k$ |
| $\alpha_m$ | Learning rate (weight) for the $m$-th tree |
| *Distributions and Densities* | |
| $f_{X_j}(\cdot\|\boldsymbol{x}_{-j})$ | Conditional probability density function (PDF) of $X_j$ |
| $F_{X_j}(\cdot\|\boldsymbol{x}_{-j})$ | Conditional cumulative distribution function (CDF) of $X_j$ |
| $\Phi(\cdot)$ | CDF of the standard normal distribution $N(0, 1)$ |

Table S5: Summary of mathematical notation and symbols used in the methodology.
